# SARS-CoV-2 vaccine Alpha and Delta variant breakthrough infections are rare and mild, but happen relative early after vaccination

**DOI:** 10.1101/2021.12.23.21268324

**Authors:** Jelissa Katharina Peter, Fanny Wegner, Severin Gsponer, Fabrice Helfenstein, Tim Roloff, Rahel Tarnutzer, Kerstin Grosheintz, Moritz Back, Carla Schaubhut, Sabina Wagner, Helena Seth-Smith, Patrick Scotton, Maurice Redondo, Christiane Beckmann, Tanja Stadler, Andrea Salzmann, Henriette Kurth, Karoline Leuzinger, Stefano Bassetti, Roland Binggisser, Martin Siegemund, Maja Weisser, Manuel Battegay, Sarah Tschudin Sutter, Aitana Lebrand, Hans H. Hirsch, Simon Fuchs, Adrian Egli

## Abstract

**Introduction:** COVID-19 vaccines significantly reduce SARS-CoV-2 (SCoV2)-related hospitalization and mortality in randomized controlled clinical trials, as well as in real-world effectiveness against different circulating SCoV2-lineages. However, some vaccine recipients show breakthrough infection and it remains unknown, which host and viral factors contribute to this risk and how many resulted in severe outcomes. Our aim was to identify demographic and clinical risk factors for SCoV2 breakthrough infections and severe disease in fully vaccinated individuals and to compare patient characteristics in breakthrough infections caused by SCoV2 Alpha or Delta variant.

**Methods:** We conducted an exploratory retrospective case-control study from 28th of December to 25th of October 2021 dominated by the Delta SCoV2 variant. All cases of infection had to be reported by law to the local health authorities. Vaccine recipients’ data was anonymously available from the national Vaccination Monitoring Data Lake and the main local vaccine center. We compared anonymized patients’ characteristics of breakthrough infection (n=492) to two overlapping control groups including all vaccine recipients from the Canton of Basel-City (group 1 n=126’586 and group 2 n=109’382). We also compared patients with breakthrough infection caused by the Alpha to Delta variant. We used different multivariate generalized linear models (GLM).

**Results:** We found only 492/126’586 (0.39%) vaccine recipients with a breakthrough infection after vaccination during the 10 months observational period. Most cases were asymptomatic or mild (478/492 97.2%) and only very few required hospitalization (14/492, 2.8%). The time to a positive SCoV2 test shows that most breakthrough infections occurred between a few days to about 170 days after full vaccination, with a median of 78 days (interquartile range, IQR 47-124 days). Factors associated with a lower odds for breakthrough infection were: age (OR 0.987, 95%CI 0.983-0.992), previous COVID-19 infection prior to vaccination (OR 0.296, 95%CI 0.117-0.606), and (self-declared) serious side-effects from previous vaccines (OR 0.289, 95%CI 0.033-1.035). Factors associated with a higher odds for breakthrough infection were: vaccination with the Pfizer/BioNTech vaccine (OR 1.459, 95%CI 1.238-1.612), chronic disease as vaccine indication (OR 2.109, 95%CI 1.692-2.620), and healthcare workers (OR 1.404, 95%CI 1.042-1.860). We did not observe a significantly increased risk for immunosuppressed patients (OR 1.248, 95% CI 0.806-1.849).

**Conclusions:** Our study shows that breakthrough infections are rare and show mild illness, but that it occurs early after vaccination with more than 50% of cases within 70 to 80 days post-full vaccination. This clearly implies that boost vaccination should be much earlier initiated compared to the currently communicated 180-day threshold. This has important implications especially for risk groups associated with more frequent breakthrough infections such as healthcare workers, and people in high-risk care facilities. Due to changes in the epidemiological dynamic with new variants emerging, continuous monitoring of breakthrough infections is helpful to provide evidence on booster vaccines and patient groups at risk for potential complications.

## Introduction

The COVID-19 vaccines induce SARS-CoV-2 (SCoV2)-specific neutralizing antibodies in most vaccine recipients ^1-4^ and have shown significant efficacy to reduce COVID-19 related hospitalization and mortality in randomized controlled clinical trials ^5-7^, as well as in real-world effectiveness against different circulating SCoV2 variants ^8-10^. However, some vaccine recipients show breakthrough infection either shortly or longer after vaccination ^5,11-13^. It is not yet fully understood, which factors contribute to lower vaccine effectiveness ^14,15^ and how to best identify them for a more personalized vaccination and booster strategy. In short-term breakthrough infections, potential reasons include IgA deficiency, immunosuppressive drugs e.g., after organ transplantation ^16-18^, or immunosenescence ^19^. In long-term breakthrough infections, most likely SARS-CoV-2 specific neutralizing antibodies titers slowly decrease over time and fall below a certain threshold, which results in an increasing risk of a breakthrough infection ^20^. For this reason, additional third or even fourth vaccine boosters have become clinical practice in many countries by which neutralization titers can be elevated. The World Health Organisation (WHO) recently published an interim statement on booster doses, stating that additional vaccination at standard doses or boosters at reduced doses should be firmly evidence-driven and targeted to the population groups in greatest need ^21^.

Beside important host related factors, viral evolution may also contribute to the risk of breakthrough infections, as observed during previous influenza seasons ^22^. Constantly emerging new SCoV2 variants may reduce vaccine efficacy and effectiveness by modulating the viral spike (S)-protein in the receptor binding domain (RBD) ^23,24^, which is critical for binding to the angiotensin converting enzyme (ACE)-2 receptor on the host cell ^25-27^. Since the beginning of the SCoV2 pandemic, certain mutations in the viral S-protein have been retained and spread globally, whereby Delta variant (B.1.617.2) has been rapidly displacing most of the other circulating viral variants in Europe. It has been shown that the efficiency to neutralize the Delta variant is lower compared to wild type or Alpha variants. Two-times vaccinated individuals showed a 2.5-fold (Pfizer/BioNtech, Moderna and Janssen vaccine) reduction in neutralization titres against the Delta virus compared to an ancestral G614 S pseudo-virus ^28^ and a 3-fold (Pfizer/BioNtech) and 6-fold (AstraZeneca) reduction in neutralization titres against the Alpha variant ^24^. The rapid molecular epidemiological adaptation and recent introduction of the Delta and Omicron SCoV2 variants makes it more difficult to conclusively define risk factors for breakthrough infection ^29,30^. Recent data shows that the new Omicron variant may show a 40-fold reduction to be neutralized in double dose vaccinated people ^31^.

Currently, there are two key questions of high importance for public health decision making regarding vaccination and booster application. Firstly, what are possible demographic and clinical risk factors for SCoV2 breakthrough infections in fully vaccinated individuals? Knowledge would allow us to make general recommendations and prioritize third dose applications. Second, do patient characteristics differ in breakthrough infections caused by SCoV2 Alpha or Delta variant? The answer could help to link viral- and host-factors risk assessments and learn for the emerging Omicron variant.

Since the beginning of the Swiss COVID-19 vaccination campaign in December 2020, for which only uses licensed vaccines from Moderna or Pfizer/BioNTech, we have documented all breakthrough infections of the Canton of Basel-City, including those occurring in persons having received other vaccines licensed in other countries such as AstraZeneca and Janssen. We noted an increasing number of breakthrough infections in fall 2021. Therefore, we conducted an exploratory retrospective case-control study and aimed to identify factors potentially associated with breakthrough infections in fully vaccinated people. We use a fully anonymized dataset of more than 120’000 fully vaccinated residents of the Canton of Basel-City, who received two doses of the licensed SCoV2 vaccines. We then investigated potential differences in breakthrough infections with either the Alpha or the Delta SCoV2 variants.

## Methods

### Ethics

The study has been approved by the local ethical committee (EKNZ 2020-00769 and 2021-00774). The data protection office of the Canton of Basel-City also evaluated and approved the study. We received fully anonymized data with minimal demographic information such as age, gender, and self-reported data to conduct the study. For cases of breakthrough infections, we had minimal clinical information available based on the mandatory reporting system and contact tracing information. This data was also anonymized and different teams in data acquisition and analysis were involved. No identification of single individuals is possible.

### Setting

The Canton of Basel-City has three municipalities with a total population of 201’386 people (June 2021) and is the most densely populated Swiss canton (5’433/km^2^) with a population density comparable to Madrid (5’481/km^2^), London (5’701/km^2^), and Boston (5’397/km^2^) ^32-35^. The City of Basel itself is inhabited by 86% of the canton’s population. The first COVID-19 vaccine in the Canton of Basel-City was administered on the 28th of December 2020, only 20 days after the launch of the global vaccination program. In the supplementary material, a detailed description of the time course on vaccination timelines is available (**Supplementary Material**). By the 25th of October 2021 126’586 individuals, equalling 63% of the canton’s population (201’354 total inhabitants in October 2021 ^36^), were fully vaccinated against SARS-COV-2 with either BNT162b2 (Pfizer/BioNTech), mRNA-1273 (Moderna), or Ad26.COV2.S (Janssen).

### Study design and population

We conducted an exploratory retrospective case-control study from 28th of December 2020 to 25th of October 2021. Inclusion criteria for the *case cohort* were: (i) Individuals fully vaccinated in the Canton of Basel-City, including in retirement and nursing homes and hospitals, (ii) people living in the Canton of Basel-City: including Basel, Bettingen, and Riehen, and (iii) people with either SCoV2 specific NAT or antigen-test confirmed infection. Inclusion criteria for the *control cohort* were: (i) Individuals fully vaccinated at the cantonal vaccination center or at health care institutions, pharmacies, and hospitals; and (ii) people living in the Canton of Basel-City including Basel, Bettingen, and Riehen. The exclusion criterion for the case cohort was a positive NAT result considered as residual finding from a SARS-CoV-2 infection before the final vaccination. There were no exclusion criteria for the control group.

### Data collection

Data on all vaccinated individuals were collected from the Vaccination Monitoring Data Lake (VMDL) of Switzerland and the Corona Vaccination Centre for the Canton of Basel-City. All vaccinating sites in Switzerland are obliged to report to the national register according to the Swiss Epidemic law, with a 100% coverage of the canton’s total population. Data on positive tested individuals with a breakthrough infection were collected through the cantonal contact tracing database. All individuals with residence in the Canton of Basel-City tested positive for SCoV2 (RNA and antigen) are being reported to the Federal Office of Public Health (FOPH) and through there to the cantonal Contact Tracing. Thereafter each positive tested individual is contacted and monitored throughout the isolation, whereby personal and clinical information is being collected for source identification. For hospitalized individuals the clinical information is being collected through the attending physician or nurse. We identified two sub-control groups: control group 1 comprises individuals fully vaccinated against COVID-19, who got vaccinated in all health care institutions, pharmacies and hospitals, including the vaccine center Basel city. Control group 2 is a subgroup of control group 1 and comprises individuals fully vaccinated against COVID-19 in the Corona Vaccination Centre for the Canton of Basel-City. Although meant as a control set, both control groups also comprise individuals who suffered from a breakthrough infection. Anonymization prevents tracking back individual information and thus prevents the exclusion of individuals with a breakthrough infection from the control sets.

Vaccine recipient characteristics have been collected as self-reported variables and were used to describe cases, to explain the probability of undergoing a breakthrough infection or to explain the probability of undergoing a breakthrough infection caused by either the Alpha or Delta SCoV2 variants. Supplementary table 1 shows an overview on control groups and patients with breakthrough infections (**Supplementary Table 1**).

The clinical course of infection was classified according to the National Institute of Health (NIH) COVID-19 treatment guidelines into “asymptomatic infection” (no symptoms), “mild illness” (various signs e.g., fever, cough; but no shortness of breath, dyspnea or abnormal chest imaging), “moderate illness” (evidence of lower respiratory disease and oxygen saturation (SpO2) ≥94% on room air at sea level), “severe illness” (SpO2 <94% on room air at sea level, PaO2/FiO2 <300 mm Hg, respiratory frequency <30 breaths/min, or lung infiltrates >50%), “critical illness” (respiratory failure, septic shock, and/or multiple organ dysfunction) and “death” ^37^.

### SCoV2 lineage determination

At the University Hospital Basel, we used viral whole genome sequencing to determine the lineage in cases identified as breakthrough infection by Health Services of the Canton of Basel-City. Briefly, we used the Arctic protocol on a NextSeq 500 platform (Illumina), followed by the GOVGAP pipeline to determine the viral lineage as previously described ^38,39^. In a subset of samples the sequencing was done in another laboratory. These sequences could be accessed via the Swiss Pathogen Surveillance Platform (www.spsp.ch) and ENA and GISAID accessions. The accession numbers of the genomes used for this analysis are available in the supplementary table 7 (**Supplementary Table 2**).

### Full vaccination

Full vaccination was defined as (a) seven days after the second BNT162b2 vaccine (Pfizer/BioNTech) or (b) seven days after the first BNT162b2 vaccine when the patient had a confirmed SCoV2 infection at least 30 days before the dose, (c) 14 days after the second mRNA-1273 (Moderna) vaccine or (d) 14 days after the first mRNA-1273 when the patient had a confirmed SARS-CoV-2 infection at least 30 days before the dose, (e) 14 days after a second ChAdOx1-S vaccine (AstraZeneca) and (f) 22 days after a single dose of Ad26.COV2.S (Janssen).

### Breakthrough infection

A breakthrough infection was defined by a laboratory-confirmed SCoV2 infection through either SCoV2-specific nucleic acid testing (NAT) or antigen testing in fully vaccinated individuals. In Switzerland, positive antigen tests are recommended to be confirmed by NAT to exclude the rare possibility of false-positive results whenever possible, thereby also providing access to samples for SCoV2 genome sequencing. The date of the breakthrough infection was set to the date of sample collection.

### Statistical analysis

The analysis set is the full analysis set obtained by merging the case group with the control group 1 and 2 respectively. The data to be used in this study is information routinely collected at the time of vaccination or at the time of consultation in case of breakthrough infection. Missing values were not imputed. Hence statistical analyses were based on available case sets. All analyses were performed with R version 4.0.3 (2020-10-10). All variables were explored as a descriptive analysis including median and interquartile ranges of continuous variables unless otherwise indicated and as numbers and percentage for categorical variables.

Four models were used. The reported statistics for all models included: Odds-ratios, confidence intervals and associated p-values. Model 1: The analysis was conducted using control individuals from both group 1 and 2 respectively and case patients. The risk factors were included into a multivariate generalized linear model (GLM) with logit link-function and binomial distribution (logistic model) to investigate their potential association with the probability of suffering from a breakthrough infection or not. Model 2: In order to investigate whether the two control subgroups represented two different populations and whether the same or different risk factors are associated with the probability of a breakthrough infection when using control group 1 or 2, we ran two additional multivariate GLMs including the same risk factors but using either the dataset comprising control individuals from group 1 and case patients or the dataset comprising control individuals from group 2 and case patients. Model 3: In order to investigate whether severe immunosuppression further explains the probability of undergoing a breakthrough, we ran another multivariate GLM including all risk factors used in the previous models plus severe immunosuppression. This analysis was carried out on a dataset comprising control individuals from control group 2 and case patients. Model 4: To investigate which, if any, risk factors are associated with the probability of undergoing a breakthrough infection caused by the Alpha or Delta SCoV2 variants, we conducted a multivariate GLM with logit link-function and binomial distribution including the case group risk factors.

Although p-values were provided, readers should be aware that these cannot be used to accept or reject the null or the alternative hypothesis based on some predefined thresholds. P-values assess the compatibility of the data with the null hypothesis on a continuous scale. The data interpretation is not based on an arbitrary dichotomous threshold nor on p-values alone, but by combining p-values with a thorough examination of the effect size (odds-ratio) and considering the amount of uncertainty (error) around it (95% confidence interval).

## Results

### Occurrence of breakthrough infections

We identified a total of 492 patients (0.39%) with breakthrough infections in an overall population of 126’586 vaccine recipients. First, we analyzed the time distribution, in days, elapsed from full vaccination to the positive SCoV2 test result for all patients with breakthrough infections. The time to a positive SCoV2 test shows that most breakthrough infections occurred between a few days to about 170 days after full vaccination, with a median of 78 days (interquartile range, IQR 47-124 days). The data suggests a weak bi-modal distribution, with a much weaker peak after 180 days. Accordingly, a steady increase of breakthrough infections starting after about 10-20 days post-vaccine and gradually levelling off after about 130 days in the frequency and cumulative frequency distributions (**Figure 1**). The burden of breakthrough infections correlated with the occurrence of overall SCoV2 cases in the observed community (**Supplementary Figure 1**).

**Figure 1.**
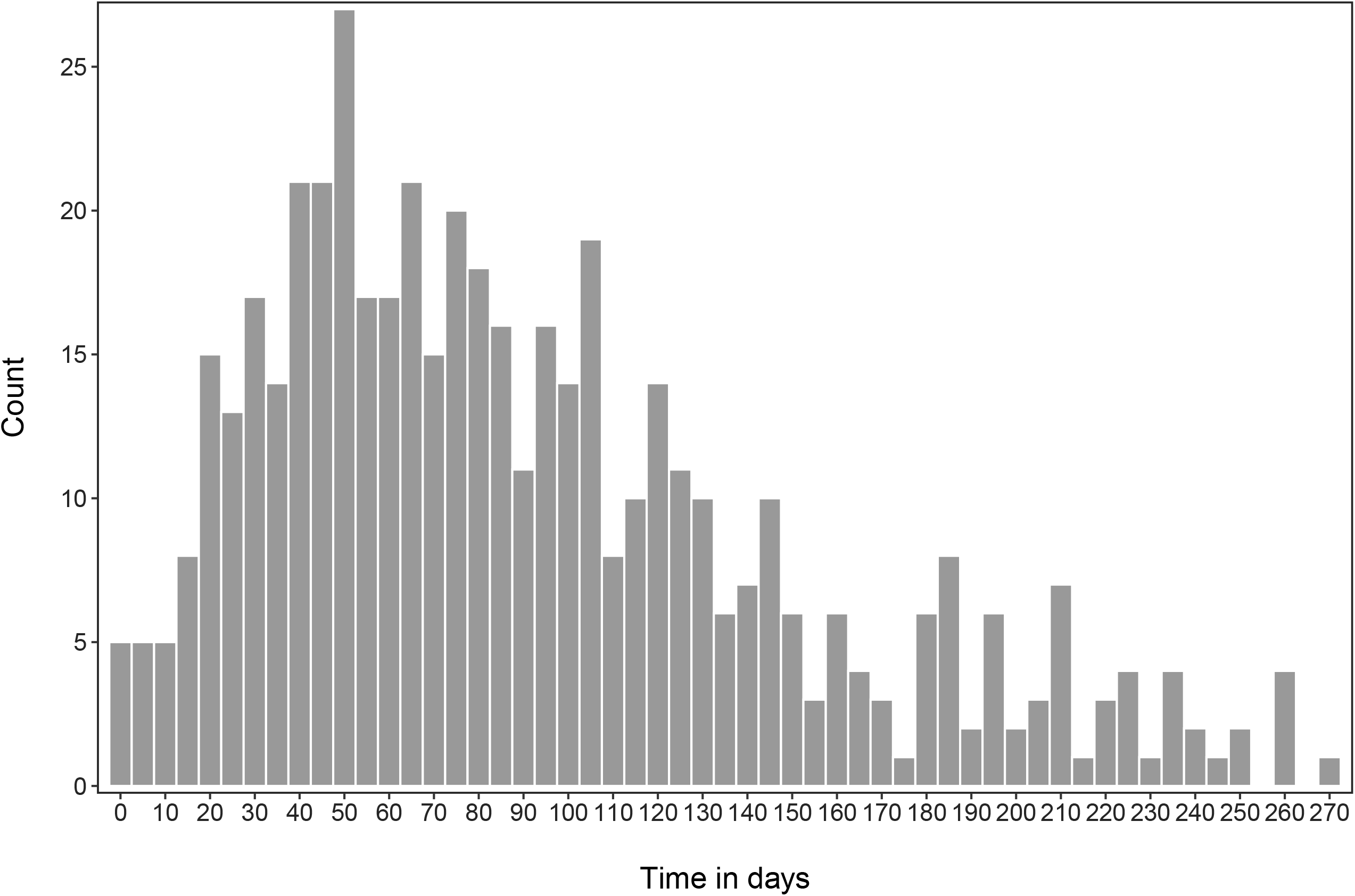
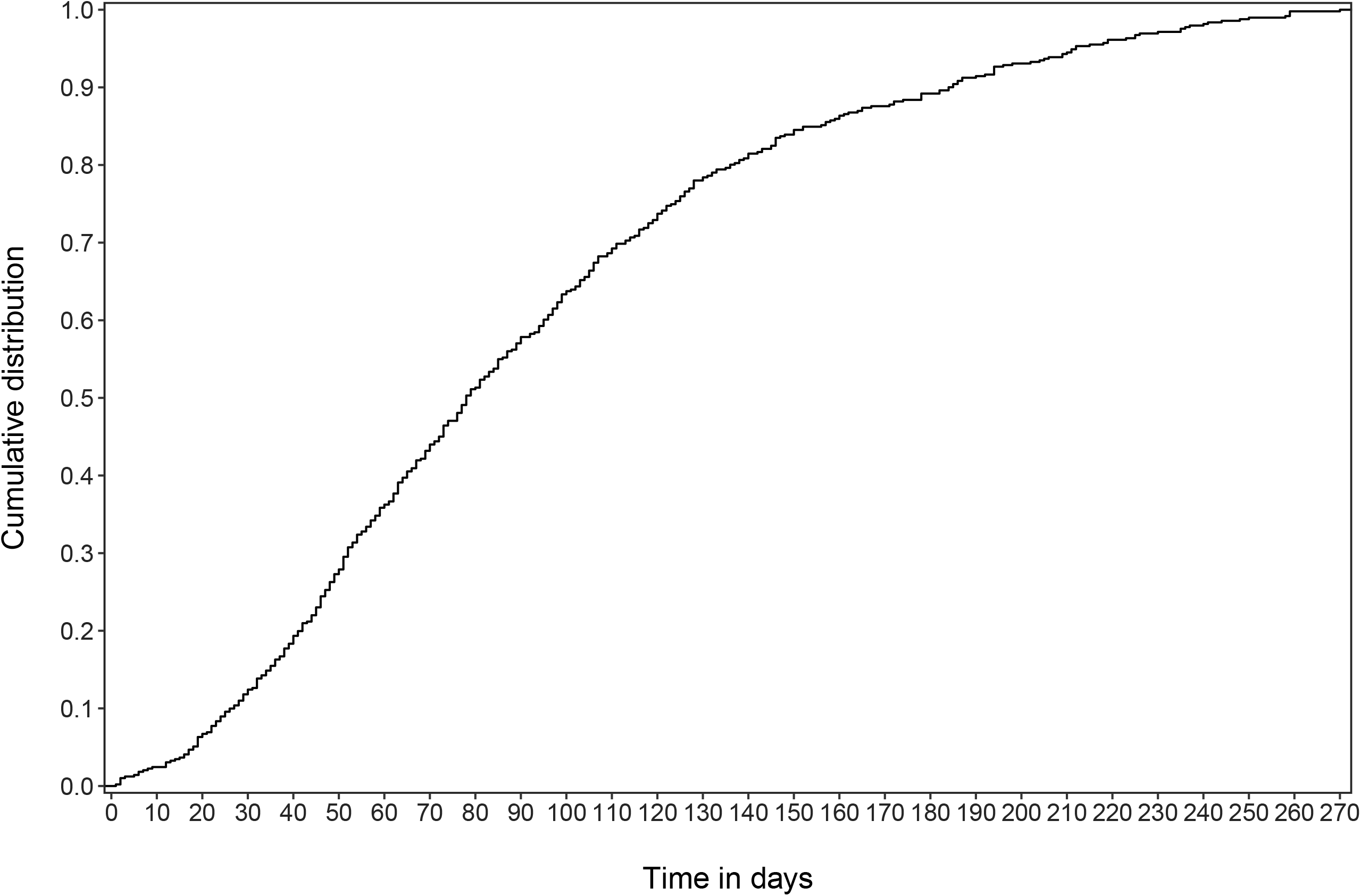
Distribution time in days from full vaccination (as defined above) to the positive SCoV2 test result confirming all cases of breakthrough infection. Upper panel, absolute frequency; lower panel, cumulative frequency.

### Clinical case presentation

We further analyzed all available patient characteristics for vaccine recipients, who suffered from a breakthrough infection (**Supplementary Table 3**). Briefly, patients with a breakthrough infection had a median age of 45 years (IQR 32-64) and were slightly more often female (52.6%). The mRNA-1273 (Moderna) and BNT162b2 mRNA (Pfizer/BioNTech) were the two most commonly used COVID19 vaccines in our region, accounting for 99.2% of vaccine applications. Generally, people with breakthrough infections had an asymptomatic presentation or mild illness (26/492, 5.3% and 452/492, 91.9%). Only a minority of patients required hospitalization (14/492, 2.8%). Severe illness (5/492, 1.0%) and critical illness or death (2/492, 0.2%) were very rare. Patients requiring hospitalization were generally older compared to patients who did not require hospitalization (median age 82 years vs. 44 years). Hospitalized patients also showed more common predisposing risk-factors compared to non-hospitalized patients e.g., cardiac disease 64.3% vs 6.5% and arterial hypertension 50% vs. 14% (**Supplementary Table 4**). Similarly, patients who showed moderate, severe and critical illness also showed higher age (77, 81, and 87 years) and were having more predisposing risk factors compared to patients with asymptomatic or mild disease e.g., cardiac disease 71.4%, 80%, and 100% (**Supplementary Table 5**). In a subset of patients, we could also determine the Alpha and Delta SCoV2 variants. The Delta variant showed more breakthrough infections compared to the Alpha variant in absolute numbers. Of note, this reflected the epidemiological situation in the general population during the observational period, where the Delta variant was the dominant virus lineage in Fall 2021.

Next, we compared patients with breakthrough infections to all vaccine recipients using the two control groups: control group 1 (n=126’586) and 2 (n=109’382) (**Table 1**). The median age in patients with SCoV2 breakthrough infection was slightly lower compared to the control group 1 and 2 (median 45 years vs. 49 years vs. 52 years). The patients with SCoV2 breakthrough infections were more frequently vaccinated with the Pfizer/BioNTech vaccine compared to the control group 1 and 2 (47% vs. 33.5% vs. 33.1%). Also notable is that people at especially high risk according to the Federal Office of Public Health e.g., with a chronic disease (detailed list of conditions in **Supplementary Table 6**, which is a key indication for vaccination) suffered more frequently from breakthrough infection than the control group 1 and 2 (31.9% vs. 20.5% vs. 22.4%). Next, we compared characteristics, which were only available in patients with a breakthrough infection and people of control group 2 (which is a subset of control group 1). We observed slightly more breakthrough infections in patients with severe immunosuppression (5.3% vs. 3.0%) and less people who had serious side-effects after previous vaccines (0.2% vs. 1.0%) (**Table 1**).

**Table 1.**
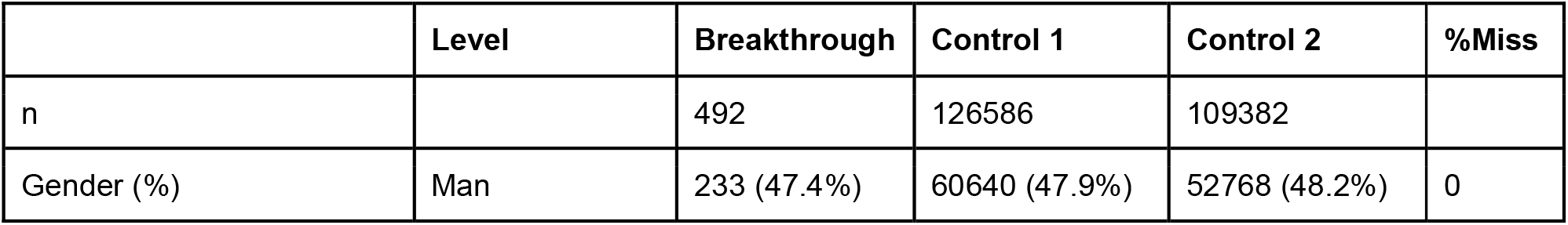
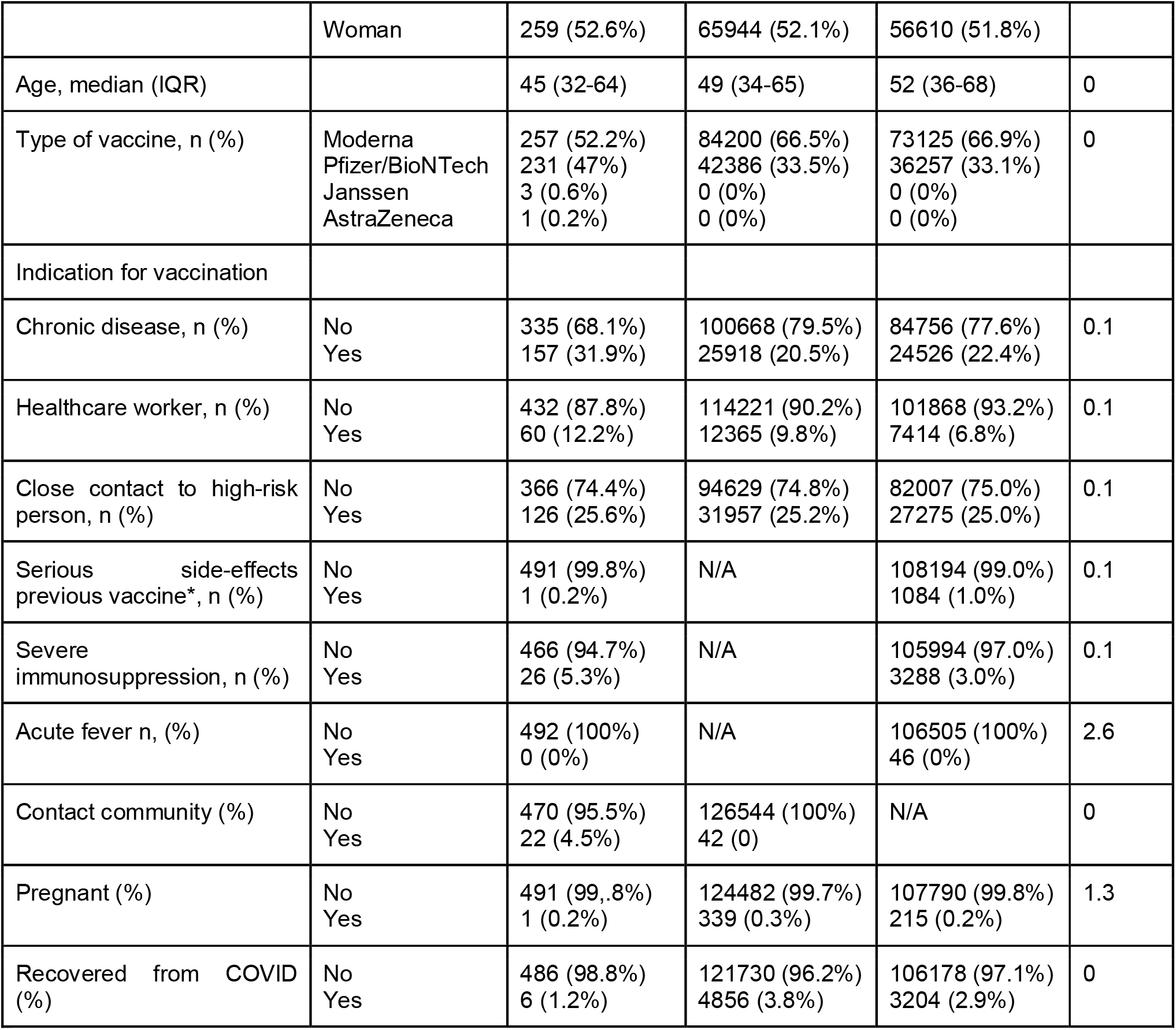
Comparison of patients with breakthrough infections compared to all control vaccine recipients using the control sets. Categorical variables are presented with numbers and percentages. *Serious side-effects in previous vaccines focuses on any vaccine and is a selfreported variable. Since age shows some departure from normality in the case group, it is presented with median and [Interquartile range].

|  | Level | Breakthrough | Control 1 | Control 2 | %Miss |
| --- | --- | --- | --- | --- | --- |
| n |  | 492 | 126586 | 109382 |  |
| Gender (%) | Man | 233 (47.4%) | 60640 (47.9%) | 52768 (48.2%) | 0 |
|  | Woman | 259 (52.6%) | 65944 (52.1%) | 56610 (51.8%) |  |
| Age, median (IQR) |  | 45 (32-64) | 49 (34-65) | 52 (36-68) | 0 |
| Type of vaccine, n (%) | Moderna<br>Pfizer/BioNTech<br>Janssen<br>AstraZeneca | 257 (52.2%)<br>231 (47%)<br>3 (0.6%)<br>1 (0.2%) | 84200 (66.5%)<br>42386 (33.5%)<br>0 (0%)<br>0 (0%) | 73125 (66.9%)<br>36257 (33.1%)<br>0 (0%)<br>0 (0%) | 0 |
| Indication for vaccination |  |  |  |  |  |
| Chronic disease, n (%) | No<br>Yes | 335 (68.1%)<br>157 (31.9%) | 100668 (79.5%)<br>25918 (20.5%) | 84756 (77.6%)<br>24526 (22.4%) | 0.1 |
| Healthcare worker, n (%) | No<br>Yes | 432 (87.8%)<br>60 (12.2%) | 114221 (90.2%)<br>12365 (9.8%) | 101868 (93.2%)<br>7414 (6.8%) | 0.1 |
| Close contact to high-risk person, n (%) | No<br>Yes | 366 (74.4%)<br>126 (25.6%) | 94629 (74.8%)<br>31957 (25.2%) | 82007 (75.0%)<br>27275 (25.0%) | 0.1 |
| Serious side-effects previous vaccine*, n (%) | No<br>Yes | 491 (99.8%)<br>1 (0.2%) | N/A | 108194 (99.0%)<br>1084 (1.0%) | 0.1 |
| Severe immunosuppression, n (%) | No<br>Yes | 466 (94.7%)<br>26 (5.3%) | N/A | 105994 (97.0%)<br>3288 (3.0%) | 0.1 |
| Acute fever n, (%) | No<br>Yes | 492 (100%)<br>0 (0%) | N/A | 106505 (100%)<br>46 (0%) | 2.6 |
| Contact community (%) | No<br>Yes | 470 (95.5%)<br>22 (4.5%) | 126544 (100%)<br>42 (0) | N/A | 0 |
| Pregnant (%) | No<br>Yes | 491 (99.8%)<br>1 (0.2%) | 124482 (99.7%)<br>339 (0.3%) | 107790 (99.8%)<br>215 (0.2%) | 1.3 |
| Recovered from COVID (%) | No<br>Yes | 486 (98.8%)<br>6 (1.2%) | 121730 (96.2%)<br>4856 (3.8%) | 106178 (97.1%)<br>3204 (2.9%) | 0 |

### Identification of potential risk factors

Next, we identified potential risk factors related to the probability of suffering from a breakthrough infection using generalized linear models. We compared patients with a breakthrough infection towards control group 1 (**Table 2**) and control group 2 (**Supplementary Table 7**). Overall, both multivariate regression models including control group 1 and 2 showed comparable results.

**Table 2.** Multivariable logistic regression exploring the relationships between potential risk factors and the probability to suffer from a SCoV2 breakthrough infection using the ‘control’ set 1. Only Moderna and Pfizer/BioNTech vaccines are compared, because no other vaccine was used in the control population. A generalized linear mixed model was used.

|  | n | Odds ratio | CI | p |
| --- | --- | --- | --- | --- |
| Intercept | 125308 | 0.006 | (0.005-0.008) | <0.001 |
| Age |  | 0.987 | (0.983-0.992) | <0.001 |
| Gender (woman vs. man) |  | 1.037 | (0.867-1.242) | 0.692 |
| Vaccine (Pfizer/BioNTech, ref =Moderna) |  | 1.459 | (1.238-1.612) | <0.001 |
| Chronic disease (=Yes) |  | 2.109 | (1.692-2.620) | <0.001 |
| Healthcare worker (=Yes) |  | 1.404 | (1.042-1.860) | 0.022 |
| Close contact with high-risk person (=Yes) |  | 0.902 | (0.724-1.116) | 0.349 |
| Pregnant (=Yes) |  | 0.671 | (0.038-2.995) | 0.691 |
| Recovered from COVID-19 (=Yes) |  | 0.296 | (0.117-0.606) | 0.003 |

A higher age was associated with a lower odds for breakthrough infections (OR 0.987, 95% CI 0.983-0.992). Therefore, we further explored, if breakthrough infections were differently distributed between the age quartiles. Whereas 16 to 64 years old patients showed breakthrough infections mainly from July to October 2021, corresponding to infections caused by the Delta SCoV2 variant, patients above 64 years old showed a first bulk of breakthrough infections from January to May 2021, corresponding to infections caused by the alpha variant, and a second, more important bulk from July to October corresponding to infections caused by the Delta variant (**Figure 2**), suggesting a different epidemiology as younger age groups could not get the vaccine as early, hence no breakthrough infection was possible.

**Figure 2.**
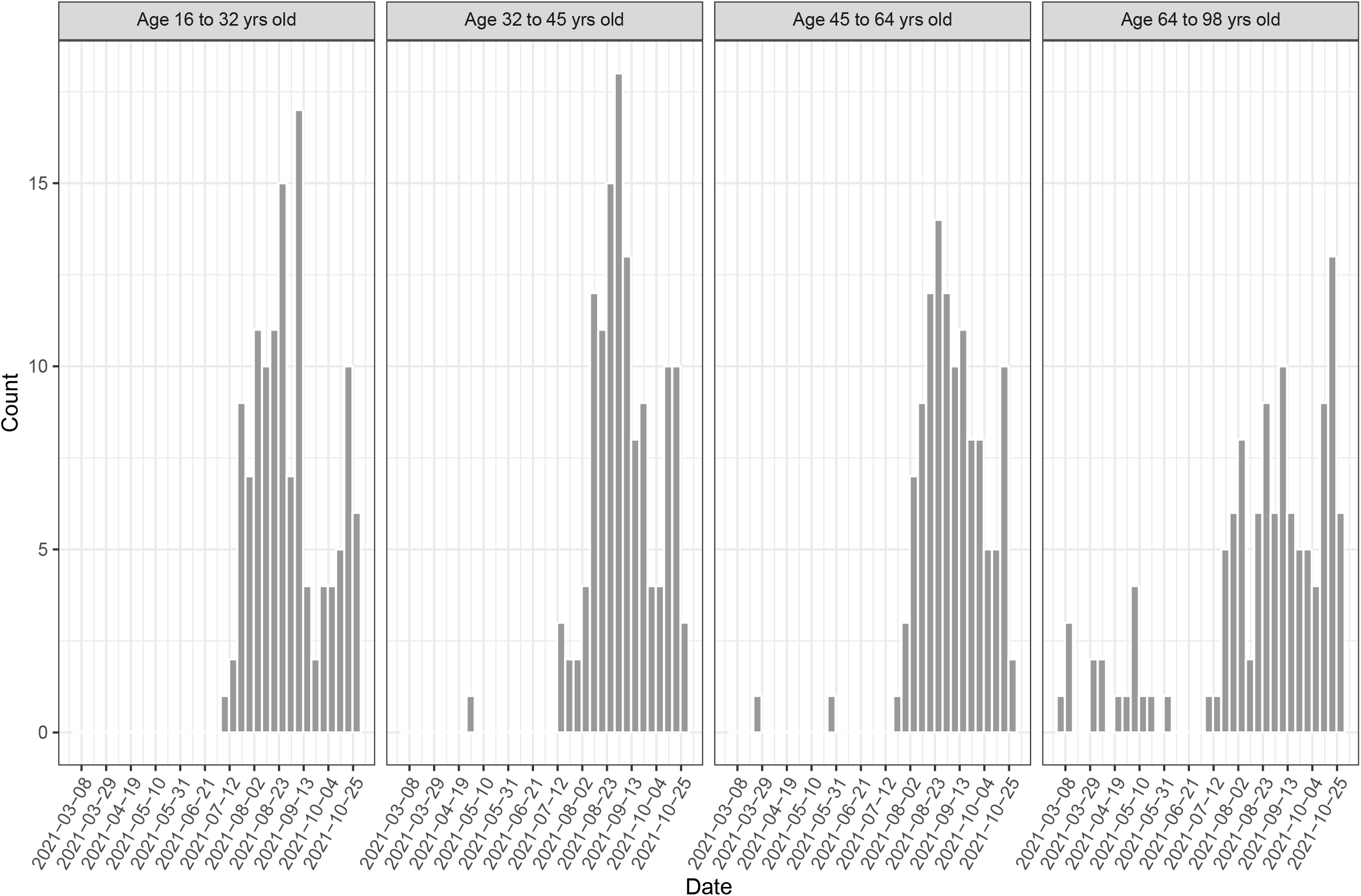
Age quartiles and distribution of breakthrough infections.

Breakthrough infections were more common in people vaccinated with the Pfizer/BioNTech vaccine (OR 1.459), with chronic disease (OR 2.109) and in healthcare workers (OR 1.404). Patients who recovered from COVID-19 prior to vaccination were less likely to suffer from a breakthrough infection (OR 0.296). We did not observe a significantly increased risk for immunosuppressed patients (OR 1.248, 95% CI 0.806-1.849). Patients with (self-declared) serious side-effects from previous vaccines show a slightly lower risk for breakthrough infection (OR 0.289, 95% CI 0.033-1.035) (**Supplementary Table 8**). We also performed a subgroup analysis focusing on breakthrough infection with the Delta variant (**Supplementary Table 9 and 10**).

### Difference between breakthrough infections with Alpha or Delta SCoV2 variants

Next, we explored potential differences between breakthrough infections caused by the Alpha or Delta SCoV2 variants (**Table 3**). Supplementary Figure 2 and 3 show detailed time distributions for breakthrough infection caused by these variants (**Supplementary Figure 2 and 3**). Based on the Swiss national surveillance program for SCoV2 and shared data efforts of the Swiss Pathogen Surveillance Platform (www.spsp.ch), we know that during Fall 2021 about 97% of the circulating viruses in Switzerland were Delta variants and subtypes thereof (https://www.covid19.admin.ch/en/). The sequences of the viral isolates identified are part of normal phylogenetic distribution and do not show specific unusual mutations e.g., in the viral Spike protein (data not shown). Of note, during the observational period we have not observed the newly discovered Omicron variant (B.1.529). Currently this variant rapidly increases and accounts for approximately 7% of cases in Switzerland (as of 10th December 2021, https://www.covid19.admin.ch/en/).

**Table 3.** Comparisons of patient characteristics between individuals who suffered from a breakthrough infection caused by the Alpha or the Delta variant. Categorical variables are presented with numbers and percentages. *Serious side-effects in previous vaccines focuses on any vaccine and is a self-reported variable. Variables showing departure from normality are presented with median and [Interquartile Range].

| Variable | Level | SCoV2-Alpha | SCoV2-Delta | % missing |
| --- | --- | --- | --- | --- |
| n |  | 12 | 258 |  |
| Age, med (IQR) | Years | 82.0 (70.5-87.0) | 44.0 (32.0-61.8) | 45.1 |
| Sex, n (%) | Man<br>Woman | 5 (41.7%)<br>7 (58.3%) | 124 (48.1%)<br>134 (51.9%) | 45.1 |
| Type of vaccine, n (%) | Moderna<br>Pfizer/BioNTech<br>Janssen<br>AstraZeneca | 1 (8.3%)<br>11 (91.7%)<br>0 (0%)<br>0 (0%) | 136 (52.7%)<br>120 (46.5%)<br>1 (0.4%)<br>1 (0.4%) | 45.1 |
| Indication for vaccination |  |  |  |  |
| Chronic diseases, n (%) | No<br>Yes | 1 (8.3%)<br>11 (91.7%) | 183 (70.9%)<br>75 (29.1%) | 45.1 |
| Healthcare worker, n (%) | No<br>Yes | 12 (100%)<br>0 (0%) | 222 (86.0%)<br>36 (14.0%) | 45.1 |
| Close contact to high-risk person, n (%) | No<br>Yes | 6 (50%)<br>6 (50%) | 191 (74.0%)<br>67 (26.0%) | 45.1 |
| Serious side effects to other vaccines*, n (%) | No<br>Yes | 12 (100%)<br>0 (0%) | 257 (99.6%)<br>1 (0.4%) | 45.1 |
| Severe immunosuppression, n (%) | No<br>Yes | 9 (75%)<br>3 (25%) | 247 (95.7%)<br>11 (4.3%) | 45.1 |
| Acute fever, n (%) | No<br>Yes | 12 (100%)<br>0 (0%) | 258 (100%)<br>0 (0%) | 45.1 |
| Pregnant, n (%) | No<br>Yes | 12 (100%)<br>0 (0%) | 257 (99.6%)<br>1 (0.4%) | 45.1 |
| Recovered from COVID19, n (%) | No<br>Yes | 12 (100%)<br>0 (0%) | 256 (99.2%)<br>2 (0.8%) | 45.1 |
| Time vacc to 1st symptoms, median (IQR) | days | 59 (57-61) | 75 (44-118) | 45.1 |
| Time vacc/Delta outbreak to 1st symptoms, median (IQR) | days | 59 (57-61) | 49.5 (28.3-72.0) | 45.1 |
| Time vacc to 1st symptoms for Alpha & Delta patients vaccinated after Delta outbreak, median (IQR) | days | 59 (57-61) | 40 (23-57) | 45.1 |
| Time vacc to positive test, median (IQR) | days | 42 (17-63) | 77 (46-120) | 45.1 |
| Time vacc/Delta outbreak to positive test, median (IQR) | days | 42 (17-63) | 49 (31-72) | 45.1 |
| Time vacc to positive test for Alpha & Delta patients vaccinated after Delta outbreak, median (IQR) | days | 42 (17-63) | 45 (26-59) | 45.1 |
| PCR ct value, median (IQR) |  | 20 (20-20) | 20.02 (17.22-23.18) | 45.1 |
| Severity of disease, n (%) | Asymptomatic<br>Mild illness<br>Moderate illness<br>Severe illness<br>Critical illness and death | 2 (16.7%)<br>7 (58.3%)<br>0 (0%)<br>2 (16.7%)<br>1 (8.3%) | 6 (2.3%)<br>244 (94.6%)<br>5 (1.9%)<br>2 (0.8%)<br>1 (0.4%) | 45.1 |
| Hospitalisation | No<br>Yes | 11 (91.7%)<br>1 (8.3%) | 127 (96.9%)<br>8 (3.1%) | 45.1 |
| Source of infection | Club, party, bar<br>Cultural event<br>Family<br>Friends<br>Holidays<br>Hospital<br>Religion<br>Retirement and nursing home<br>School<br>Sports<br>Unknown<br>Work | 0 (0%)<br>0 (0%)<br>3 (25%)<br>0 (0%)<br>1 (8.3%)<br>1 (8.3%)<br>0 (0%)<br>5 (41%)<br>0 (0%)<br>0 (0%)<br>2 (16.7%)<br>0 (0%) | 12 (4.7%)<br>4 (1.6%)<br>72 (27.9%)<br>14 (5.4%)<br>63 (24.4%)<br>3 (1.2%)<br>1 (0.4)<br>10 (3.9%)<br>1 (0.4%)<br>3 (1.2%)<br>58 (22.5%)<br>17 (6.6%) | 45.1 |
| Baseline risk factor |  |  |  |  |
| Chronic lung disease | No<br>Yes | 11 (91.7%)<br>1 (8.3%) | 243 (94.2%)<br>15 (5.8%) | 45.1 |
| Chronic renal disease | No<br>Yes | 11 (91.7%)<br>1 (8.3%) | 254 (98.4%)<br>4 (1.6%) | 45.1 |
| Cancer | No<br>Yes | 10 (83.3%)<br>2 (16.7%) | 253 (98.1%)<br>5 (1.9%) | 45.1 |
| Cardiac disease | No<br>Yes | 7 (58.3%)<br>5 (41.7%) | 240 (93.0%)<br>18 (7.0%) | 45.1 |
| Arterial hypertension | No<br>Yes | 9 (75%)<br>3(25%) | 215 (83.3%)<br>43 (16.7%) | 45.1 |
| Diabetes | No<br>Yes | 10 (83.3%)<br>2 (16.7%) | 234 (90.7%)<br>24 (9.3%) | 45.1 |
| Obesity | No<br>Yes | 11 (91.7%)<br>1 (8.3%) | 257 (99.6%)<br>1 (0.4%) | 45.1 |

The Alpha variant was dominating from February to June 2021 and then replaced by the Delta variant in May 2021. During this time also the vaccination concept and availability changed by providing access to individuals without high-risk disease and younger people. No difference regarding the time to positive test could be observed between the two variants. However, the two variants showed very different baseline characteristics. The median age for patients with breakthrough infection caused by the Alpha variant was 82 years and for the Delta variant only 41.5 years. This shows that breakthrough infections over time occurred in very different populations. The age difference is also reflected in the distribution of reasons for vaccination e.g., the proportion of chronic diseases (91.7% in Alpha vs. 26.2% in Delta) or living in a high-risk community facility (41.7% vs. 0.8%). Although the median time, in days, from vaccination to a confirmed SCoV2 breakthrough infection was different with 42 days in the Alpha variant compared to 69 days in the Delta variant, this difference was not significant. It also seems that case severity in Alpha variants was higher compared to Delta variants, e.g., severe and critical illnesses accounted for 25% (Alpha) and 1.6% (Delta), which could also reflect the same issue of a more vulnerable population. For the Alpha variant, breakthrough infection was also more often linked to retirement and nursing homes (41%), whereas for the Delta variant mainly linked to holidays and work (23.1% and 6.9%).

Supplementary Table 8 summarizes the results of a generalized linear model with binomial distribution investigating potential risk factors associated with the relative probability of being infected by the Delta variant (vs. Alpha variant taken as a reference) in case of breakthrough infection (**Supplementary Table 8**).

## Discussion

Our study explores the key factors associated with COVID19 vaccine breakthrough infection and compares the patient characteristics between people with a breakthrough infection caused by either the Alpha or Delta SCoV2 variant. The key findings of our study are: (i) breakthrough infections are very rare with only 0.39% (492/126586) of the vaccinated population being affected during a 10 months observation period; (ii) breakthrough infections were either asymptomatic or mild (97.2%) with only 2.8% of patients (14/492) requiring hospitalization (14/126586; 0.01%); (iii) most of the breakthrough infection in our population occurred between a few days to about 170 days after full vaccination, with a median of only 78 days for the time to a positive SCoV2 test.

In our cohort, older individuals are less likely to suffer from a breakthrough infection. This may be due to younger individuals being more likely to travel, enjoy social events outside their home and thus be more exposed to the Delta variant, which caused most of the breakthrough infections in our study ^40,41^. Especially after vaccination, young and healthy individuals might feel secure, leading to a higher risk behavior. Similarly, individuals with close contact to a high-risk person were less likely to suffer from a breakthrough infection. This finding is most probably explained due to the more vigilant behavior as well and underlines the fact that incautious behavior might be a greater risk factor for breakthrough infection than intrinsic risk factors such as age or chronic disease.

We also observed that individuals who recovered from a previous COVID-19 infection are less likely to suffer from a breakthrough infection. This is of interest particularly because the previous infection was caused by the Alpha variant whereas the breakthrough infection was caused by the Delta SCoV2 variant in this patient subgroup. The natural infection may build different types of immune response and additional immunological memory against other viral proteins ^42,43^, recently termed hybrid immunity ^44^. Further studies regarding breakthrough infections in COVID-19 recovered patients will show whether a previous infection with the Alpha or Delta SARS-CoV-2 variant leads to sufficient protection against the emerging Omicron virus variant.

Our study indicates that individuals with a chronic disease are more likely to suffer from a breakthrough infection. Similar findings have been made mainly for the elderly with overall more chronic diseases ^14,45,46^.

Individuals vaccinated with the Pfizer/BioNTech vaccine (BNT162b2) are more likely to suffer from a breakthrough infection than individuals vaccinated with the Moderna vaccine (mRNA-1273). Tenforde and colleagues also showed that the mRNA-1273 vaccine was significantly more protective (OR, 0.11) compared with the BNT162b2 vaccine (OR, 0.19). The protective association against hospitalization for the BNT162b2 vaccine more than 120 days following vaccination declined notably (OR, 0.36; median, 143 days from vaccine dose 2 to illness onset), whereas the effectiveness of the mRNA-1273 vaccine more than 120 days post-vaccination was largely preserved (OR, 0.15; median, 141 days from vaccine dose 2 to illness onset) ^6^. Our study provides important confirmation and points out that risk groups for more severe clinical outcomes may benefit from the Moderna vaccine, due to the reduction in breakthrough infections.

Of particular relevance for the emerging new wave of Omicron is our observation that healthcare workers are more likely to suffer from a breakthrough infection. This is an important finding and highlights the importance for booster vaccines in this cohort. A recent publication showed 39 cases of breakthrough infection in a total 1497 healthcare workers, which was particularly linked to titers of SARS-CoV-2 neutralizing antibodies ^5^ and another study identified 4 of 1388 (0.3%) breakthrough infections in fully vaccinated compared to 21 of 585 (3.6%) in non-vaccinated staff of a long-term care facility. Most likely the increased risk of breakthrough infection in healthcare workers is explained through a higher exposure to infected people.

Further, in our cohort, most breakthrough infections were caused by the Delta variant. However, interpreting the temporal dynamics of infection relative to vaccination for the Alpha and Delta variant is difficult since these variants emerged at different time points relative to the availability of vaccination in Switzerland. Consequently, the median time to positive test does not necessarily demonstrate the ability of the vaccine to protect individuals from a SCoV2 infection, but is probably biased by the variant itself and the non-pharmaceutical public health measures during the course of the study. Our additional models focussing only on individuals vaccinated after the emergence of the Delta variant, show a median time of 38 days (IQR 17-54) to SCoV2-positive testing. As the subgroup analysis resulted in a loss of statistical power, the cumulative distributions no longer can resolve whether or not a breakthrough infection is more likely to occur as more time elapsed since full vaccination. Other studies have reported an increasing risk with a longer time since vaccination ^20^. In our vaccine control cohort, the median time since vaccination was 115 days (IQR 84-145, min=0, max=273) and 82 days (IQR 51-100, min=0, max=111) for individuals vaccinated after the Delta outbreak. The hypothesis of breakthrough infections increasing with elapsed time since full vaccination must consider new circulating SCoV2 variants and the overall increase in case numbers during this period. Indeed, our results are consistent with the notion that individuals infected with the Delta variant were at higher risk for breakthrough infection compared to individuals infected with the Alpha variant. Although, we have not determined prospectively neutralization antibody titers, this phenomenon is most likely explained by the amino acid changes on the virus spike protein and subsequently the resulting lower neutralization efficiency ^24,28^ and the general higher number of infected individuals with the Delta variant.

Surveillance of viral evolution using whole genome sequencing on a national scale remains critical for the months to come. As shown in our data as well, the clinical course of vaccine breakthrough infection has been described as generally mild (https://www.cdc.gov/coronavirus/2019-ncov/vaccines/effectiveness/why-measure-effectiveness/breakthrough-cases.html), with a few exceptions ^47^. It is therefore crucial to also closely monitor the frequency and severity of SARS-CoV-2 breakthrough infections, including the clinical course, and combine this information with surveillance of viral evolution associated with reducing vaccine effectiveness, and their clinical course in fully vaccinated individuals with breakthrough infection. This has been done in retrospect but based on reported databases real-time monitoring of breakthrough infection should be initiated.

### Limitations

Our study has several important limitations. During the study timeframe, diagnostic and screening tests on SARS-CoV-2 for vaccinated individuals were only recommended by the Federal Office of Public Health (FOPH) when symptoms occurred or when the vaccinated individual lived in the same household as an individual tested positive for SARS-CoV-2. Therefore, although all positive test results are being reported to the FOPH, missing data on breakthrough infections in e.g., asymptomatic patients is possible. Due to different reporting systems, information regarding immunosuppression in the control population is only available for individuals vaccinated in the Corona Vaccination Centre for the Canton of Basel-City and not for individuals vaccinated in other sites such as pharmacies or hospitals. Because only vaccinating institutions in Switzerland are obligated to report to the Swiss COVID-19 vaccination register, individuals with residence in the Canton of Basel-City vaccinated outside of Switzerland are not recorded in the database. Accordingly, individuals vaccinated with ChAdOx1-S only appear in the case group. We included a relatively small number of sequenced cases with breakthrough infection - sequencing was available only for a subset of PCR confirmed cases (approximately 5-10%) of all cases are part of a nation wide surveillance program. Although we have not sequenced all strains, it is unlikely that we have missed a substantial dominant new lineage causing breakthrough infection, as the lineages dominating during the observational period were also found in other parts of the country as dominant lineages and also in the surrounding neighboring countries. Also, we do not have neutralizing antibody titers available for the whole cohort, however, this is also not a suitable assay for a general and large population.

### Conclusion

Overall our study shows that breakthrough infections are rare and mild, and that the highest risk is early after vaccination - more than 50% of breakthrough infections occur 70 to 80 days post-vaccine. This clearly implies that boost vaccination should be much earlier initiated compared to the currently communicated 180 day threshold. This has important implications especially for risk groups associated with more frequent breakthrough infections such as healthcare workers, and people in high risk care facilities. Due to changes in the epidemiological dynamic with new variants emerging, continuous monitoring of breakthrough infection may be very helpful to provide evidence on booster vaccines and patient groups at risk for potential complications.

## Data Availability

All data produced in the present work are contained in the manuscript. All genome sequences used are mentioned in Supplementary Table 2 with the respective accession numbers on GISAID.

## Acknowledgment

We thank the members of the different SARS-CoV-2 vaccination teams in Basel. We also want to thank the Swiss Pathogen Surveillance Platform (www.spsp.ch) for providing access to genomes and lineage related information of cases with breakthrough infection.

## Figure legends

**Supplementary Figure 1**. SARS-CoV-2 PCR confirmed cases after introduction of the vaccine. The red line shows all breakthrough infections and the black line shows all PCR confirmed cases (secondary axis) in Basel-City.

**Supplementary Figure 2**. Distribution time in days from full vaccination (as defined) to the breakthrough infections with the Alpha variant. Upper panel, absolute frequency; lower panel cumulative frequency.

**Supplementary Figure 3**. Distribution time in days from full vaccination (as defined) to the breakthrough infections with the Delta variant. Upper panel, absolute frequency; lower panel cumulative frequency.

Supplementary Figure 3.

**Supplementary Table 1.** Collected vaccine recipient and patient characteristics. * other reasons than age, healthcare worker, chronic disease, contact with vulnerable person, living in a community with increased risks. * general previous side effects.

| Control group 1 | Control group 2 | Breakthrough infections |
| --- | --- | --- |
| Age |  | Age |
| Gender, (male/female), count & percentage | Gender, (male/female), count & percentage | Gender, (male/female), count & percentage |
| Type of vaccine; categorical, Pfizer/BioNTech, Moderna, Janssen, AstraZeneca; count & percentage | Type of vaccine; categorical, Pfizer/BioNTech, Moderna, Janssen, AstraZeneca; count & percentage | Type of vaccine; categorical, Pfizer/BioNTech, Moderna, Janssen, AstraZeneca; count & percentage |
| Chronic disease, (Yes/No) | Chronic disease, (Yes/No) | Chronic disease, (Yes/No) |
| Healthcare worker, (Yes/No) | Healthcare worker, (Yes/No) | Healthcare worker, (Yes/No) |
| Close contact to high-risk person, (Yes/No) | Close contact to high-risk person, (Yes/No) | Close contact to high-risk person, (Yes/No) |
| Vaccination for other reasons*, (Yes/No) | N/A |  |
| Pregnancy, (Yes/No) | Pregnancy, (Yes/No) | Pregnancy, (Yes/No) |
| Person recovered from COVID-19, (Yes/No) | Person recovered from COVID-19, (Yes/No) | Person recovered from COVID-19, (Yes/No) |
| Date of full vaccination | Date of full vaccination | Date of full vaccination |
| N/A | Severe immunosuppression, (Yes/No) | Severe immunosuppression, (Yes/No) |
| N/A | Serious previous side effects*, (Yes/No) | Serious side effects, (Yes/No) |
| N/A | Acute fever before / during COVID-19 vaccination, (Yes/No) | Acute fever before / during COVID-19 vaccination, (Yes/No) |
| N/A | N/A | Time between full vaccination and positive test |
| N/A | N/A | Time between full vaccination and first symptoms |
| N/A | N/A | SARS-CoV-2 variant, (Alpha or Delta), count & percent |
| N/A | N/A | Hospitalisation, (Yes/No) |
| N/A | N/A | Suspected source of infection: club, party, bar; cultural event; family; friends; holidays; hospital; religion; retirement and nursing home; school; sports; work; unknown; |
| N/A | N/A | Chronic lung disease, (Yes/No) |
| N/A | N/A | Chronic renal disease, (Yes/No) |
| N/A | N/A | Cancer, (Yes/No) |
| N/A | N/A | Cardiovascular disease, (Yes/No) |
| N/A | N/A | Arterial hypertension, (Yes/No) |
| N/A | N/A | Diabetes, (Yes/No) |
| N/A | N/A | Obesity, (Yes/No) |

**Supplementary Table 2.** Accession numbers of used genomes in this analysis.

| **GISAID Identifier** | **Pangolin Lineage** |
| --- | --- |
| EPI_ISL_3086290 | AY.4 |
| EPI_ISL_4903597 | AY.4 |
| EPI_ISL_4903635 | AY.102 |
| EPI_ISL_5049804 | AY.102 |
| EPI_ISL_5307606 | AY.46 |
| EPI_ISL_5307608 | AY.43 |
| EPI_ISL_5307610 | AY.54 |
| EPI_ISL_5307620 | AY.43 |
| EPI_ISL_5307622 | AY.14 |
| EPI_ISL_5307667 | AY.46 |
| EPI_ISL_5307709 | AY.102 |
| EPI_ISL_5428846 | AY.4 |
| EPI_ISL_5428847 | AY.41 |
| EPI_ISL_5428848 | AY.4 |
| EPI_ISL_5428849 | AY.46.6 |
| EPI_ISL_5428850 | AY.111 |
| EPI_ISL_5508919 | AY.43 |
| EPI_ISL_5508977 | AY.98.1 |
| EPI_ISL_5508986 | AY.98.1 |
| EPI_ISL_5508994 | AY.43 |
| EPI_ISL_5509063 | AY.80 |
| EPI_ISL_5509099 | AY.47 |
| EPI_ISL_5509125 | AY.33 |
| EPI_ISL_5509131 | AY.33 |
| EPI_ISL_5509160 | AY.43 |
| EPI_ISL_5509166 | AY.4 |
| EPI_ISL_5765992 | AY.43 |
| EPI_ISL_5766012 | AY.95 |
| EPI_ISL_5766062 | AY.43 |
| EPI_ISL_5766070 | AY.43 |
| EPI_ISL_5766077 | AY.43 |
| EPI_ISL_5766144 | AY.43 |
| EPI_ISL_5766182 | B.1.617.2 |
| EPI_ISL_5766183 | AY.20 |
| EPI_ISL_5766195 | B.1.617.2 |
| EPI_ISL_5766203 | AY.43 |
| EPI_ISL_5766208 | AY.43 |
| EPI_ISL_5766269 | AY.46.6 |
| EPI_ISL_5841634 | AY.44 |
| EPI_ISL_5841635 | AY.54 |
| EPI_ISL_5841657 | AY.4 |
| EPI_ISL_5841661 | AY.102 |
| EPI_ISL_5841666 | AY.43 |
| EPI_ISL_5841685 | AY.4 |
| EPI_ISL_5841691 | AY.102 |
| EPI_ISL_5841709 | AY.9.2 |
| EPI_ISL_5933964 | AY.102 |
| EPI_ISL_5933970 | AY.102 |
| EPI_ISL_5934004 | AY.4 |
| EPI_ISL_5934005 | AY.102 |
| EPI_ISL_5934006 | AY.43 |
| EPI_ISL_5934007 | AY.43 |
| EPI_ISL_5934008 | AY.4 |
| EPI_ISL_5934009 | AY.102 |
| EPI_ISL_5934010 | AY.46.6 |
| EPI_ISL_6015872 | AY.44 |
| EPI_ISL_6015873 | AY.43 |
| EPI_ISL_6015874 | AY.43 |
| EPI_ISL_6015875 | AY.43 |
| EPI_ISL_6157569 | AY.54 |
| EPI_ISL_6157570 | AY.102 |
| EPI_ISL_3086299 | AY.70 |
| EPI_ISL_5841659 | AY.43 |
| EPI_ISL_1747622 | B.1.1.7 |
| EPI_ISL_1747636 | B.1.1.7 |
| EPI_ISL_1747641 | B.1.1.7 |
| EPI_ISL_1748263 | B.1.1.7 |
| EPI_ISL_1748463 | B.1.1.7 |
| EPI_ISL_1914607 | B.1.1.7 |
| EPI_ISL_2270095 | B.1.1.7 |
| EPI_ISL_2466583 | B.1.1.7 |
| EPI_ISL_3030104 | AY.98.1 |
| EPI_ISL_3030113 | AY.43 |
| EPI_ISL_3086318 | AY.42 |
| EPI_ISL_3086319 | B.1.617.2 |
| EPI_ISL_3184351 | AY.43 |
| EPI_ISL_3184366 | AY.9.2 |
| EPI_ISL_3229188 | B.1.1.7 |
| EPI_ISL_3229190 | AY.43 |
| EPI_ISL_3229201 | AY.43 |
| EPI_ISL_3229213 | B.1.617.2 |
| EPI_ISL_3229214 | B.1.617.2 |
| EPI_ISL_3229224 | AY.5 |
| EPI_ISL_3229225 | AY.43 |
| EPI_ISL_3229227 | AY.43 |
| EPI_ISL_3229248 | AY.4 |
| EPI_ISL_3247301 | AY.43 |
| EPI_ISL_3247308 | AY.42 |
| EPI_ISL_3247349 | AY.98.1 |
| EPI_ISL_3303743 | AY.43 |
| EPI_ISL_3303758 | AY.42 |
| EPI_ISL_3344075 | AY.42 |
| EPI_ISL_3344101 | AY.4 |
| EPI_ISL_3344102 | AY.43 |
| EPI_ISL_3344111 | B.1.617.2 |
| EPI_ISL_3425805 | AY.98.1 |
| EPI_ISL_3425806 | AY.34 |
| EPI_ISL_3451674 | AY.43 |
| EPI_ISL_3451732 | AY.43 |
| EPI_ISL_3451733 | AY.43 |
| EPI_ISL_3530135 | AY.42 |
| EPI_ISL_3530171 | AY.94 |
| EPI_ISL_3530179 | AY.23 |
| EPI_ISL_3530776 | AY.43 |
| EPI_ISL_3530792 | AY.98.1 |
| EPI_ISL_3530842 | AY.34 |
| EPI_ISL_3570114 | AY.16 |
| EPI_ISL_3570115 | B.1.617.2 |
| EPI_ISL_3570127 | AY.43 |
| EPI_ISL_3570128 | B.1.617.2 |
| EPI_ISL_3570130 | AY.43 |
| EPI_ISL_3710254 | AY.4 |
| EPI_ISL_3710255 | AY.98.1 |
| EPI_ISL_3730669 | AZ.2 |
| EPI_ISL_3730670 | AY.36 |
| EPI_ISL_3824224 | AY.42 |
| EPI_ISL_3824225 | AY.4 |
| EPI_ISL_3824226 | AY.98.1 |
| EPI_ISL_3824227 | AZ.2 |
| EPI_ISL_3824228 | AY.43 |
| EPI_ISL_3824229 | AY.43 |
| EPI_ISL_3824230 | AY.102 |
| EPI_ISL_3824270 | AY.12 |
| EPI_ISL_4251188 | AY.43 |
| EPI_ISL_4251192 | AY.43 |
| EPI_ISL_4251219 | AY.43 |
| EPI_ISL_4392868 | AY.43 |
| EPI_ISL_4395046 | AY.9.1 |
| EPI_ISL_4395061 | AY.43 |
| EPI_ISL_4395072 | AY.4 |
| EPI_ISL_4395085 | AY.104 |
| EPI_ISL_4395102 | AY.86 |
| EPI_ISL_4395109 | AY.43 |
| EPI_ISL_4395150 | AY.33 |
| EPI_ISL_4395151 | AY.9.2 |
| EPI_ISL_4395152 | AY.102 |
| EPI_ISL_4395153 | AY.9.2 |
| EPI_ISL_4395154 | AY.43 |
| EPI_ISL_4395155 | AY.42 |
| EPI_ISL_4395156 | AY.98.1 |
| EPI_ISL_4395157 | AY.4 |
| EPI_ISL_4395158 | AY.43 |
| EPI_ISL_4395159 | AY.4 |
| EPI_ISL_4395160 | B.1.617.2 |
| EPI_ISL_4395161 | B.1.617.2 |
| EPI_ISL_4395162 | B.1.617.2 |
| EPI_ISL_4395163 | AY.9.2 |
| EPI_ISL_4395165 | AY.9.2 |
| EPI_ISL_4395166 | AY.43 |
| EPI_ISL_4395167 | AY.9.2 |
| EPI_ISL_4395168 | AY.43 |
| EPI_ISL_4395169 | AY.46 |
| EPI_ISL_4395170 | AY.9.2 |
| EPI_ISL_4395173 | AY.9.2 |
| EPI_ISL_4395288 | AY.5 |
| EPI_ISL_4395290 | AY.5.4 |
| EPI_ISL_4395301 | AY.73 |
| EPI_ISL_4395303 | AY.43 |
| EPI_ISL_4395308 | AY.25 |
| EPI_ISL_4410796 | AY.43 |
| EPI_ISL_4410804 | AY.9.2 |
| EPI_ISL_4410806 | AY.4 |
| EPI_ISL_4410862 | AY.42 |
| EPI_ISL_4410869 | AY.46.6 |
| EPI_ISL_4410890 | AY.4 |
| EPI_ISL_4450796 | AY.102 |
| EPI_ISL_4450798 | B.1.617.2 |
| EPI_ISL_4450800 | AY.43 |
| EPI_ISL_4450829 | AY.42 |
| EPI_ISL_4450830 | AY.9.2 |
| EPI_ISL_4450831 | AY.9.2 |
| EPI_ISL_4450832 | B.1.617.2 |
| EPI_ISL_4450833 | AY.4 |
| EPI_ISL_4450834 | AY.4 |
| EPI_ISL_4450835 | AY.43 |
| EPI_ISL_4450836 | AY.98.1 |
| EPI_ISL_4450837 | AY.46.6 |
| EPI_ISL_4450838 | AY.46.6 |
| EPI_ISL_4450839 | AY.43 |
| EPI_ISL_4450840 | AY.9.2 |
| EPI_ISL_4450841 | AY.98.1 |
| EPI_ISL_4450888 | AY.42 |
| EPI_ISL_4450889 | AY.43 |
| EPI_ISL_4450892 | AY.33 |
| EPI_ISL_4450897 | AY.43 |
| EPI_ISL_4450901 | AY.43 |
| EPI_ISL_4450904 | AY.43 |
| EPI_ISL_4542104 | AY.25 |
| EPI_ISL_4542107 | AY.39 |
| EPI_ISL_4542117 | AY.23 |
| EPI_ISL_4542185 | AY.43 |
| EPI_ISL_4571409 | B.1.617.2 |
| EPI_ISL_4571425 | AY.43 |
| EPI_ISL_4814044 | B.1.617.2 |
| EPI_ISL_4814050 | AY.102 |
| EPI_ISL_4814097 | AY.43 |
| EPI_ISL_4814115 | AY.43 |
| EPI_ISL_4903661 | AY.102 |
| EPI_ISL_4903668 | AY.43 |
| EPI_ISL_5307621 | B.1.617.2 |
| EPI_ISL_5307655 | AY.43 |
| EPI_ISL_5307682 | B.1.617.2 |
| EPI_ISL_5307686 | AY.4.3 |
| EPI_ISL_5307719 | AY.43 |
| EPI_ISL_5307727 | AY.111 |
| EPI_ISL_5307728 | AY.46 |
| EPI_ISL_5307730 | AY.43 |
| EPI_ISL_5509155 | AY.43 |
| EPI_ISL_5766178 | AY.44 |
| EPI_ISL_6307802 | AY.4 |
| EPI_ISL_6307803 | B.1.617.2 |
| EPI_ISL_6307804 | AY.25 |
| EPI_ISL_6307805 | AY.102 |
| EPI_ISL_6307806 | AY.102 |
| EPI_ISL_6307807 | AY.43 |
| EPI_ISL_6307808 | AY.4 |
| EPI_ISL_6307809 | AY.42 |
| EPI_ISL_6307810 | AY.43 |
| EPI_ISL_6307811 | AY.25 |
| EPI_ISL_6307812 | AY.102 |
| EPI_ISL_6307813 | AY.102 |

**Supplementary Table 3.** Baseline characteristics for all individuals in the database with a confirmed breakthrough infection.

| Variable | Level | Overall | % missing |
| --- | --- | --- | --- |
| n |  | 492 |  |
| Gender (%) | Male<br>Female | 233 (47.4%)<br>259 (52.6%) | 0 |
| Age, median (IQR) in years |  | 45.0 (32.00-64.00) | 0 |
| Vaccine type, n (%) | Moderna<br>Pfizer/BioNTech<br>Janssen<br>AstraZeneca | 257 (52.2%)<br>231 (47.0%)<br>3 (0.6%)<br>1 (0.2%) | 0 |
| Time full vaccine to 1st symptom, median (IQR) |  | 78 (47-122) | 7.5 |
| Time full vaccine/Delta outbreak to 1st symptoms, median (IQR) |  | 50 (29-72) |  |
| Time full vaccine to 1st symptoms for Alpha & Delta patients vaccinated after Delta outbreak, median (IQR) |  | 42 (25-59) |  |
| Time full vaccine to positive test, median (IQR) |  | 78 (47-124) | 0.8 |
| Time full vaccine/Delta outbreak to positive test, median (IQR) |  | 49 (29-72) |  |
| Time full vaccine to positive test for Alpha & Delta patients vaccinated after Delta outbreak, median (IQR) |  | 45 (25-60) |  |
| Indication for vaccine |  |  |  |
| Chronic diseases, n (%) | No<br>Yes | 335 (68.1%)<br>157 (31.9%) | 0 |
| Healthcare worker, n (%) | No<br>Yes | 432 (87.8%)<br>60 (12.2%) | 0 |
| Close contact to high-risk person, n (%) | No<br>Yes | 336 (74.4%)<br>126 (25.6%) | 0 |
| Serious side effects to other vaccines, n (%) | No<br>Yes | 491 (99.8%)<br>1 (0.2%) | 0 |
| Severe immunosuppression, n (%) | No<br>Yes | 466 (94.7%)<br>26 (5.3%) | 0 |
| Acute fever, n (%) | No<br>Yes | 492 (100%)<br>0 (0%) | 0 |
| Recovered from COVID-19 (%) | No<br>Yes | 486 (98.8%)<br>6 (1.2%) | 0 |
| Pregnant (%) | No<br>Yes | 491 (99.8%)<br>1 (0.2%) | 0 |
| Virus variant (%) | Alpha<br>Delta<br>Different or not<br>classified | 12 (2.4%)<br>258 (53.4%)<br>222 (45.1%) | 45.1 |
| PCR based Ct value, median (IQR) |  | 20.45 (17.82 - 26.20) | 60.2 |
| Clinical severity (%) | Asymptomatic<br>Mild illness<br>Moderate illness<br>Severe illness<br>Critical illness and death | 26 (5.3%)<br>452 (91.9%)<br>7 (1.4%)<br>5 (1.0%)<br>2 (0.4%) | 0 |
| Hospitalisation (%) | No<br>Yes | 478 (97.2%)<br>14 (2.8%) | 0 |
| Suspected source of infection (%) | Club, party, bar<br>Cultural event<br>Family<br>Friends<br>Holidays<br>Hospital<br>Religion<br>Retirement and nursing<br>home<br>School<br>Sports<br>Unknown<br>Work | 17 (3.5%)<br>7 (1.4%)<br>135 (27.4%)<br>26 (5.3%)<br>106 (21.5%)<br>7 (1.4%)<br>3 (0.6%)<br>23 (4.7%)<br>9 (1.8%)<br>4 (0.8%)<br>119 (24.2%)<br>36 (7.3%) | 0 |
| Baseline risk factor |  |  |  |
| Chronic lung disease (%) | No<br>Yes | 458 (93.1%)<br>34 (6.9%) | 0 |
| Chronic renal disease (%) | No<br>Yes | 484 (98.4%)<br>8 (1.6%) | 0 |
| Cancer (%) | No<br>Yes | 477 (97.0%)<br>15 (3.0%) | 0 |
| Cardiovascular disease (%) | No<br>Yes | 452 (91.9%)<br>40 (8.1%) | 0 |
| Arterial disease (%) | No<br>Yes | 417 (84.9%)<br>74 (15.1%) | 0.2 |
| Diabetes (%) | No<br>Yes | 451 (91.7%)<br>41 (8.3%) | 0 |
| Obesity (%) | No<br>Yes | 489 (99.4%)<br>3 (0.6%) | 0 |

**Supplementary Table 4.**
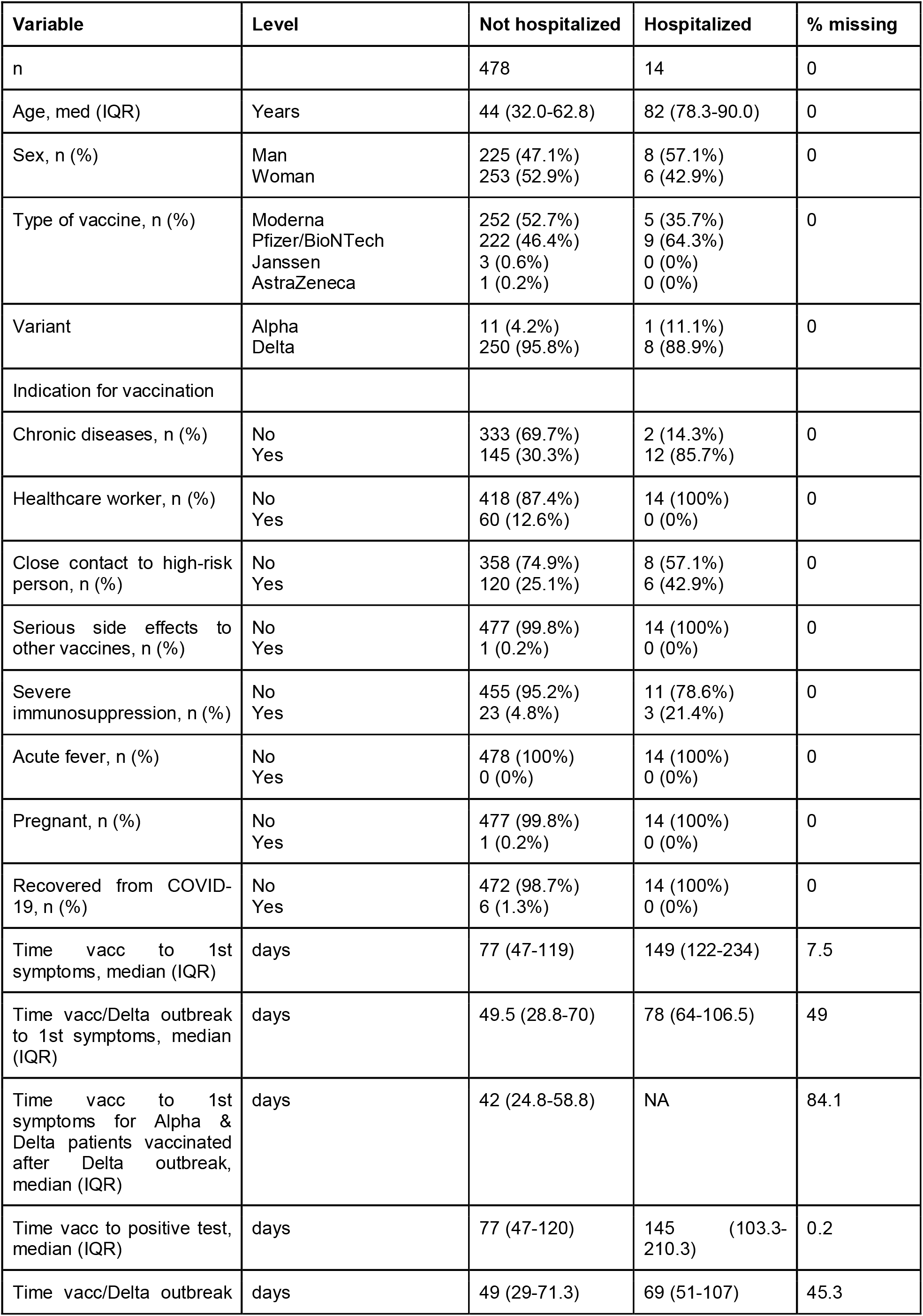
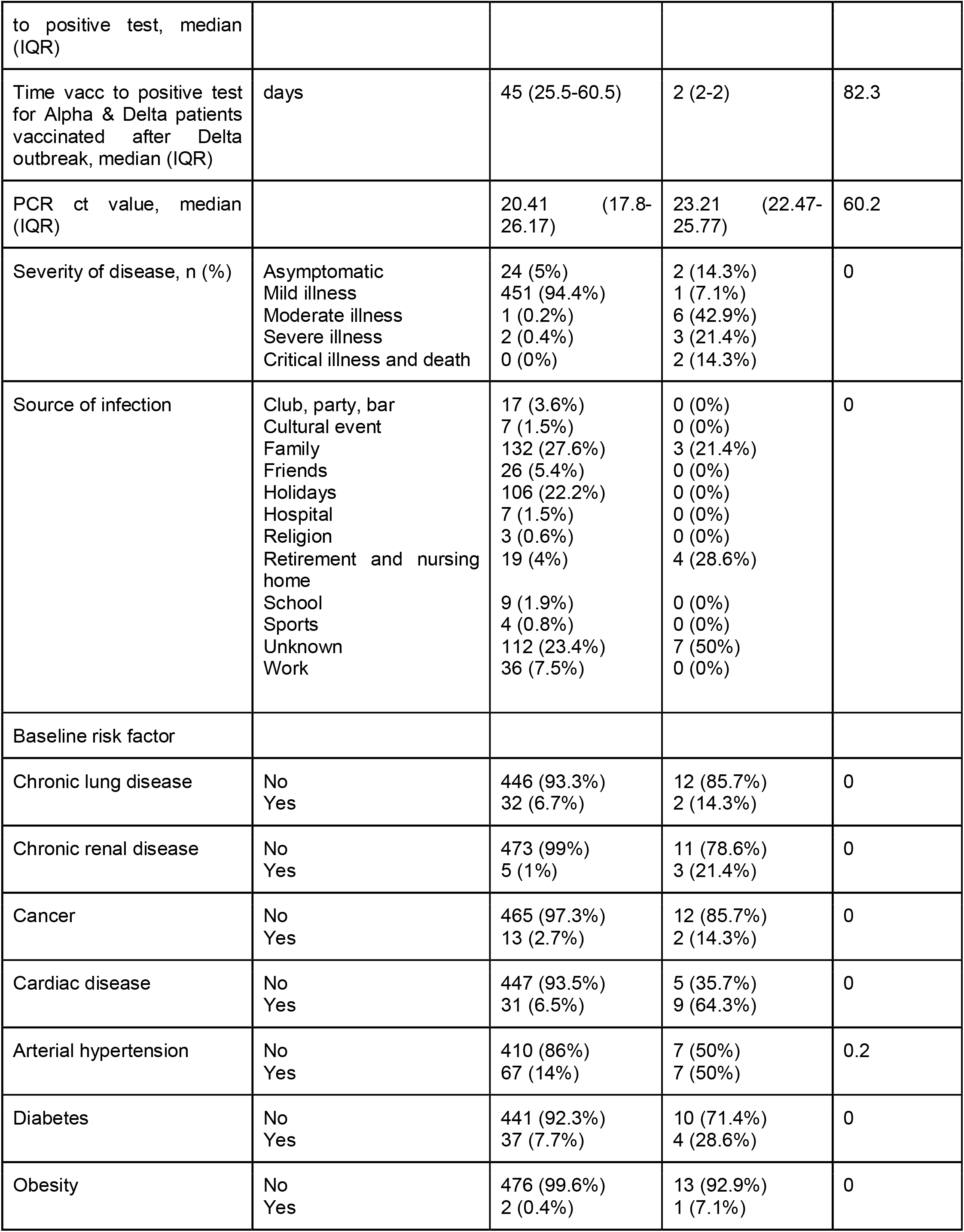
Baseline characteristics of patients with breakthrough infection who required hospitalization (n=14) compared to patients without hospitalization (n=478).

**Supplementary Table 5.** Baseline characteristics of patients with breakthrough infection who showed asymptomatic or mild illness (n=478) compared to patients with moderate, severe and critical illness (n=478).

| Variable | Level | asympto | mild | moderate | severe | critical/<br>death | %<br>miss |
| --- | --- | --- | --- | --- | --- | --- | --- |
| n |  | 26 | 452 | 7 | 5 | 2 |  |
| Age, med (IQR) | Years | 67 (32.5-82.3) | 43 (43-61.3) | 77 (63-81) | 81 (79-87) | 87 (85-89) | 0 |
| Sex, n (%) | Man<br>Woman | 10 (38.5%)<br>16 (61.5%) | 214 (47.3%)<br>238 (52.7%) | 5 (71.4%)<br>2 (28.6%) | 3 (60%)<br>2 (40%) | 1 (50%)<br>1 (50%) | 0 |
| Type of vaccine, n (%) | Moderna<br>Pfizer/BioNTech<br>Janssen<br>AstraZeneca | 13 (50%)<br>13 (50%)<br>0 (0%)<br>0 (0%) | 239 (52.9%)<br>209 (46.2%)<br>3 (0.7%)<br>1 (0.2%) | 4 (57.1%)<br>3 (42.9%)<br>0 (0%)<br>0 (0%) | 1 (20%)<br>4 (80%)<br>0 (0%)<br>0 (0%) | 0 (0%)<br>2 (100%)<br>0 (0%)<br>0 (0%) | 0 |
| Variants | Alpha<br>Delta | 2 (25%)<br>6 (75%) | 7 (2.8%)<br>244 (97.2%) | 0 (0%)<br>5 (100%) | 2 (50%)<br>2 (50%) | 1 (50%)<br>1 (50%) | 0 |
| Indication for vaccination |  |  |  |  |  |  |  |
| Chronic diseases, n (%) | No<br>Yes | 13 (50%)<br>13 (50%) | 321 (71%)<br>131 (29%) | 0 (0%)<br>7 (100%) | 1 (20%)<br>4 (80%) | 0 (0%)<br>2 (100%) | 0 |
| Healthcare worker, n (%) | No<br>Yes | 23 (88.5%)<br>3 (11.5%) | 395 (87.4%)<br>57 (12.6%) | 7 (100%)<br>0 (0%) | 5 (100%)<br>0 (0%) | 2 (100%)<br>0 (0%) | 0 |
| Close contact to high-risk person, n (%) | No<br>Yes | 14 (53.8%)<br>12 (46.2%) | 344 (76.1%)<br>108 (23.9%) | 5 (71.4%)<br>2 (28.6%) | 3 (60%)<br>2 (40%) | 0 (0%)<br>2 (100%) | 0 |
| Serious side effects to other vaccines, n (%) | No<br>Yes | 26 (100%)<br>0 (0%) | 451 (99.8%)<br>1 (0.2%) | 7 (100%)<br>0 (0%) | 5 (100%)<br>0 (0%) | 2 (100%)<br>0 (0%) | 0 |
| Severe immunosuppression, n (%) | No<br>Yes | 24 (92.3%)<br>2 (7.7%) | 431 (95.4%)<br>21 (4.6%) | 5 (71.4%)<br>2 (28.6%) | 4 (80%)<br>1 (20%) | 2 (100%)<br>0 (0%) | 0 |
| Acute fever, n (%) | No<br>Yes | 26 (100%)<br>0 (0%) | 452 (100%)<br>0 (0%) | 7 (100%)<br>0 (0%) | 5 (100%)<br>0 (0%) | 2 (100%)<br>0 (0%) | 0 |
| Pregnant, n (%) | No<br>Yes | 26 (100%)<br>0 (0%) | 451 (99.8%)<br>1 (0.2%) | 7 (100%)<br>0 (0%) | 5 (100%)<br>0 (0%) | 2 (100%)<br>0 (0%) | 0 |
| Recovered from COVID19, n (%) | No<br>Yes | 25 (96.2%)<br>1 (3.8%) | 447 (98.9%)<br>5 (1.1%) | 7 (100%)<br>0 (0%) | 5 (100%)<br>0 (0%) | 2 (100%)<br>0 (0%) | 0 |
| Time vacc to 1st symptoms, median (IQR) | days | 148 (148-148) | 77 (47-119) | 191.5 (109.3-244.5) | 123 (104-167.5) | 122 (122-122) | 7.5 |
| Time vacc/Delta outbreak to 1st symptoms, median (IQR) | days | 107 (107-107) | 49 (28.5-69.5) | 106.5 (98.3-108.3) | 64 (63-65) | 25 (25-25) | 49 |
| Time vacc to 1st symptoms for Alpha & Delta patients vaccinated after Delta outbreak, median (IQR) | days | NA | 42 (24.8-58.8) | NA | NA | NA | 84.1 |
| Time vacc to positive test, median (IQR) | days | 71.5 (41.3-135.3) | 78 (48-120.5) | 150 (96.5-242) | 85 (2-123) | 62 (32-92) | 0.2 |
| Time vacc/Delta outbreak to positive test, median (IQR) | days | 36 (20.8-57.5) | 49.5 (30.3-71.8) | 107 (78-109) | 32 (2-63.8) | 13.5 (7.8-19.3) | 45.3 |
| Time vacc to positive test for Alpha & Delta patients vaccinated after Delta outbreak, median (IQR) | days | 66.5 (44.3-88.8) | 45.5 (27.3-60.5) | NA | 2 (2-2) | 2 (2-2) | 82.3 |
| PCR ct value, median (IQR) |  | 28.8 (28.35-31.31) | 20.27 (17.73-25.74) | 25.03 (23.38-26.68) | NA | 23.21 (23.21-23.21) | 60-2 |
| Hospitalisation | No<br>Yes | 24 (92.3%)<br>2 (7.7%) | 451 (99.8%)<br>1 (0.2%) | 1 (14.3%)<br>6 (85.7%) | 2 (40%)<br>3 (60%) | 0 (0%)<br>2 (100%) | 0 |
| Source of infection | Club, party, bar<br>Cultural event<br>Family<br>Friends<br>Holidays | 1 (3.8%)<br>0 (0%)<br>5 (19.2%)<br>1 (3.8%)<br>4 (15.4%) | 16 (3.5%)<br>7 (1.5%)<br>126 (27.9%)<br>25 (5.5%)<br>102 (22.6%) | 0 (0%)<br>0 (0%)<br>4 (57.1%)<br>0 (0%)<br>0 (0%) | 0 (0%)<br>0 (0%)<br>0 (0%)<br>0 (0%)<br>0 (0%) | 0 (0%)<br>0 (0%)<br>0 (0%)<br>0 (0%)<br>0 (0%) | 0 |
|  | Hospital<br>Religion<br>Retirement and<br>nursing home<br>School<br>Sports<br>Unknown<br>Work | 0 (0%)<br>0 (0%)<br>8 (30.8%)<br>0 (0%)<br>0 (0%)<br>7 (26.9%)<br>0 (0%) | 7 (1.5%)<br>3 (0.7%)<br>11 (2.4%)<br>9 (2%)<br>4 (0.9%)<br>106 (23.5%)<br>36 (8%) | 0 (0%)<br>0 (0%)<br>1 (14.3%)<br>0 (0%)<br>0 (0%)<br>2 (28.6%)<br>0 (0%) | 0 (0%)<br>0 (0%)<br>2 (40%)<br>0 (0%)<br>0 (0%)<br>3 (60%)<br>0 (0%) | 0 (0%)<br>0 (0%)<br>1 (50%)<br>0 (0%)<br>0 (0%)<br>1 (50%)<br>0 (0%) |  |
| Baseline<br>risk factor |  |  |  |  |  |  |  |
| Chronic<br>lung<br>disease | No<br>Yes | 20 (76.9%)<br>6 (23.1%) | 426 (94.2%)<br>26 (5.8%) | 6 (85.7%)<br>1 (14.3%) | 4 (80%)<br>1 (20%) | 2 (100%)<br>0 (0%) | 0 |
| Chronic<br>renal<br>disease | No<br>Yes | 26 (100%)<br>0 (0%) | 447 (98.9%)<br>5 (1.1%) | 5 (71.4%)<br>2 (28.6%) | 5 (100%)<br>0 (0%) | 1 (50%)<br>1 (50%) | 0 |
| Cancer | No<br>Yes | 25 (96.2%)<br>1 (3.8%) | 440 (97.3%)<br>12 (2.7%) | 6 (85.7%)<br>1 (14.3%) | 4 (80%)<br>1 (20%) | 2 (100%)<br>0 (0%) | 0 |
| Cardiac<br>disease | No<br>Yes | 23 (88.5%)<br>3 (11.5%) | 426 (94.2%)<br>26 (5.8%) | 2 (28.6%)<br>5 (71.4%) | 1 (20%)<br>4 (80%) | 0 (0%)<br>2 (100%) | 0 |
| Arterial<br>hypertensio<br>n | No<br>Yes | 22 (84.6%)<br>4 (15.4%) | 388 (86%)<br>63 (14%) | 4 (57.1%)<br>3 (42.9%) | 3 (60%)<br>2 (40%) | 0 (0%)<br>2 (100%) | 0 |
| Diabetes | No<br>Yes | 25 (96.2%)<br>1 (3.8%) | 416 (92%)<br>36 (8%) | 4 (57.1%)<br>3 (42.9%) | 4 (80%)<br>1 (20%) | 2 (100%)<br>0 (0%) | 0 |
| Obesity | No<br>Yes | 26 (100%)<br>0 (0%) | 450 (99.6%)<br>2 (0.4%) | 6 (85.7%)<br>1 (14.3%) | 5 (100%)<br>0 (0%) | 2 (100%)<br>0 (0%) | 0 |

**Supplementary Table 6.** Chronic diseases and conditions leading to an especially high risk for severe course of disease according to the Federal Office of Public Health. According to the FOPH, only adults are considered to be at high risk. Therefore, the following criteria refer only to adults.

| Condition | Specific disease | Details |
| --- | --- | --- |
| Adults with Trisomy 21 |  |  |
| Hypertension | Arterial hypertension with end-organ damage |  |
|  | Therapy-resistant arterial hypertension |  |
| Cardiovascular disease | General | NYHA II-IV and NT-proBNP > 125 pg/ml |
|  |  | 2 cardio-vascular risk factors |
|  |  | Prior stroke and/or symptomatic vasculopathy |
|  |  | Chronic renal insufficiency (Stage 3, GFR <60 ml/min) |
|  | Coronary heart disease | Myocardial infarction in the past 12 months |
|  |  | Symptomatic chronic coronary syndrome despite medical treatment |
|  | Disease of the heart valves | Moderate or serious stenosis and/or regurgitation in addition to meeting at least one general criterion |
|  |  | Any surgical or percutaneous valve replacement in addition to meeting at least one general criterion |
|  | Cardiac insufficiency | Patients with dyspnea of functional class NYHA II–IV or NT-proBNP > 125pg/ml despite medical treatment for any LVEF (HFpEF, HFmrEF, HFrEF) |
|  |  | Cardiomyopathy with any cause |
|  |  | Pulmonary arterial hypertension |
|  | Arrhythmia | Auricular fibrillation with a CHA2DS2-VASc score of at least 2 points |
|  |  | Prior implant of pacemaker (incl. ICD and/or CRT implantation) in addition to meeting one general criterion |
|  | Adults with congenital heart disease | Congenital heart disease according to the individual assessment of the attending cardiologist |
| Diabetes | Diabetes mellitus, with long-term complications or a HbA1c of $\geq 8\%$ | |
| Chronic pulmonary and respiratory disease | Chronic obstructive lung diseases GOLD Grade II-IV |  |
|  | Pulmonary emphysema |  |
|  | Uncontrolled asthma, in particular serious bronchial asthma |  |
|  | Interstitial lung diseases / pulmonary fibrosis |  |
|  | Active lung cancer |  |
|  | Pulmonary arterial hypertension |  |
|  | Pulmonary vascular disease |  |
|  | Active Sarcoidosis |  |
|  | Cystic fibrosis |  |
|  | Chronic lung infections (atypical mycobacteriosis, bronchiectasis, etc.) |  |
|  | Ventilated patients |  |
|  | Diseases with severely reduced lung capacity |  |
| Diseases/Therapies that weaken the immune system | Serious immunosuppression (e.g. HIV-infection with a CD4+ T cell number of $< 200/\mu\text{l}$) | |
| | Neutropenia ($< 1'000$ neutrophils/$\mu\text{l}$) for $\geq 1$ week | |
| | Lymphocytopenia ($< 200$ lymphocytes/$\mu\text{l}$) | |
|  | Hereditary immunodeficiencies |  |
|  | Use of medication that suppresses the immune defenses (such as long-term use of glucocorticoids (prednisolone equivalent > 20 mg/day), monoclonal antibodies, cytostatics, biologics etc.) |  |
|  | Aggressive lymphomas (all entities) |  |
|  | Acute lymphatic leukaemia |  |
|  | Acute myeloid leukaemia |  |
|  | Acute promyelocytic leukaemia |  |
|  | T-cell prolymphocytic leukaemia |  |
|  | Primary lymphomas of the central nervous system |  |
|  | Stem cell transplantation |  |
|  | Amyloidosis (light-chain (AL) amyloidosis) |  |
|  | Chronic lymphatic leukaemia |  |
|  | Multiple myeloma |  |
|  | Sickle-cell disease |  |
|  | Bone marrow transplant |  |
|  | Solid organ transplant |  |
|  | Individuals on a transplant waiting list |  |
| Cancer | Cancer undergoing medical treatment |  |
| Obesity | Patients with a body-mass index (BMI) of 35 kg/m<sup>2</sup> or more |  |
| Liver disease | Cirrhosis of the liver |  |
| Renal disease | Chronic renal insufficiency with a GFR < 60 ml/min |  |

**Supplementary Table 7.** Multivariable logistic regression exploring the relationships between potential risk factors and the probability to suffer from a SARS-CoV-2 breakthrough infection using the ‘control’ group 2. Confidence intervals and p-values are computed using Firth penalization, because the level “Yes’’ for “Acute fever” was rarely observed. Only Moderna and Pfizer/BioNTech vaccines are compared, because no other vaccine was used in the control population.

|  | n | Odds ratio | CI | p |
| --- | --- | --- | --- | --- |
| Intercept | 107039 | 0.010 | (0.008-0.013) | <0.001 |
| Age |  | 0.982 | (0.977-0.987) | <0.001 |
| Gender (woman vs. man) |  | 1.048 | (0.876-1.255) | 0.606 |
| Vaccine (Pfizer/BioNTech, ref =Moderna) |  | 1.530 | (1.342-1.743) | <0.001 |
| Chronic disease (=Yes) |  | 1.990 | (1.590-2.482) | <0.001 |
| Healthcare worker (=Yes) |  | 1.927 | (1.438-2.538) | <0.001 |
| Close contact with high-risk person (=Yes) |  | 0.934 | (0.752-1.153) | 0.530 |
| Pregnant (=Yes) |  | 1.265 | (0.144-4.581) | 0.784 |
| Severe immunosuppression (=Yes) |  | 1.248 | (0.806-1.849) | 0.307 |
| Serious side-effects from previous vaccines (=Yes) |  | 0.289 | (0.033-1.035) | 0.058 |
| Acute fever (=Yes) |  | 2.693 | (0.021-19.002) | 0.549 |
| Recovered from COVID-19 (=Yes) |  | 0.431 | (0.178-0.861) | 0.014 |

**Supplementary Table 8.** Multivariable logistic regression exploring the relationships between potential risk factors and the probability to suffer from a Delta variant caused breakthrough infection using the ‘control’ set 1. Only Moderna and Pfizer/BioNTech vaccines are compared, because no other vaccine was used in the control population.

|  | n | Odds ratio | CI | p |
| --- | --- | --- | --- | --- |
| Intercept | 125076 | 0.004 | (0.003-0.006) | <0.001 |
| Age |  | 0.985 | (0.978-0.991) | <0.001 |
| Gender (woman vs. man) |  | 0.998 | (0.779-1.280) | 0.988 |
| Vaccine (Pfizer/BioNTech, ref =Moderna) |  | 1.473 | (1.232-1.759) | <0.001 |
| Chronic disease (=Yes) |  | 1.972 | (1.444-2.674) | <0.001 |
| Healthcare worker (=Yes) |  | 1.576 | (1.061-2.278) | 0.019 |
| Close contact with high-risk person (=Yes) |  | 0.918 | (0.677-1.231) | 0.575 |
| Pregnant (=Yes) |  | 1.247 | (0.071-5.614) | 0.826 |
| Recovered from COVID19 (=Yes) |  | 0.18 | (0.030-0.561) | 0.016 |

**Supplementary Table 9.** Multivariable logistic regression exploring the relationships between potential risk factors and the probability to suffer from a Delta variant caused breakthrough infection using the ‘control’ set 2. Only Moderna and Pfizer/BioNTech vaccines are compared, because no other vaccine was used in the control population.

|  | n | Odds ratio | CI | p |
| --- | --- | --- | --- | --- |
| Intercept | 106807 | 0.006 | (0.004-0.009) | <0.001 |
| Age |  | 0.979 | (0.972-0.986) | <0.001 |
| Gender (woman vs. man) |  | 1.008 | (0.787-1.293) | 0.947 |
| Vaccine (Pfizer/BioNTech, ref =Moderna) |  | 1.551 | (1.296-1.856) | <0.001 |
| Chronic disease (=Yes) |  | 1.885 | (1.374-2.568) | <0.001 |
| Healthcare worker (=Yes) |  | 2.184 | (1.484-3.126) | <0.001 |
| Close contact with high-risk person (=Yes) |  | 0.957 | (0.708-1.278) | 0.768 |
| Pregnant (=Yes) |  | 2.331 | (0.264-8.516) | 0.367 |
| Recovered from COVID-19 (=Yes) |  | 0.301 | (0.063-0.851) | 0.020 |

**Supplementary Table 10.** Multivariable generalized linear model with binomial distribution investigating the probability of suffering from a breakthrough infection caused by the Delta variant (as compared to the Alpha variant taken as the reference) as a function of individual characteristics. Confidence intervals and p-values are computed using Firth penalization due to rare or no observations in some factors’ levels.

|  | n | Odds ratio | CI | p |
| --- | --- | --- | --- | --- |
| Intercept | 270 | 977.34 | (26.01-310113.57) | <0.001 |
| Age |  | 0.93 | (0.85-0.98) | 0.007 |
| Gender (woman vs. man) |  | 1.05 | (0.21-5.39) | 0.951 |
| Vaccine Pfizer/BioNTech, (ref=Moderna) |  | 0.01 | (0.00-0.86) | 0.044 |
| Vaccine Janssen, (ref=Moderna) |  | 2.06 | (0.06-62.71) | 0.663 |
| Vaccine AstraZeneca (ref=Moderna) |  | 2.14 | (0.06-30.62) | 0.602 |
| Healthcare worker (=Yes) |  | 3.55 | (0.11-843.96) | 0.516 |
| Close contact with high-risk person (=Yes) |  | 0.62 | (0.13-2.90) | 0.534 |
| Severe immunosuppression (=Yes) |  | 0.79 | 0.05-14.70 | 0.868 |
| Pregnant (=Yes) |  | 0.01 | (0.00-4.15) | 0.116 |
| Chronic disease (=Yes) |  | 0.12 | 0.01-1.20 | 0.071 |
| Chronic lung disease (=Yes) |  | 1.37 | 0.18-22.72 | 0.780 |
| Chronic renal disease (=Yes) |  | 0.48 | 0.02-12.17 | 0.641 |
| Cancer (=Yes) |  | 0.33 | 0.01-5.07 | 0.430 |
| Cardiac disease (=Yes) |  | 0.69 | 0.10-5.12 | 0.706 |
| Arterial hypertension (=Yes) |  | 6.93 | 1.26-57.33 | 0.025 |
| Diabetes (=Yes) |  | 3.09 | 0.43-35.82 | 0.278 |
| Obesity (=Yes) |  | 0.05 | (0.00-1.16) | 0.061 |

## Supplementary Material

### Geographical setting

The City of Basel shows a population density of 7451.6/km2, a population density comparable to San Francisco (7’262.12/km2) (U.S. Census Bureau QuickFacts: San Francisco County, California) and Milan (7’681.42 ab./km2) (Demo-Geodemo. - Mappe, Popolazione, Statistiche Demografiche dell’ISTAT).

### Vaccine availability

In the beginning (January to April 2021) only elderly people above the age of 75, individuals at high risk for worse outcomes, and healthcare professionals could get vaccinated. From May 2021 onwards all Swiss citizens above the age of 16, and starting at the end of June 2021 all individuals above the age of 12 could get the vaccination. The BNT162b2 mRNA vaccine (Pfizer/BioNTech) was the first COVID-19 vaccine approved by the Swiss Agency for Therapeutic Products (Swissmedic) and therefore the only vaccine administered in December 2020. In January 2021 mRNA-1273 (Moderna) received the approval, followed by Ad26.COV2.S (Janssen) in March 2021. However, due to the missing pre-purchase of Ad26.COV2.S, the vaccine could not be administered until October 2021.

