## Supplementary figures and images for "SARS-CoV-2 vaccine Alpha and Delta variant breakthrough infections are rare and mild, but happen relative early after vaccination"

### Supplementary Figure 1

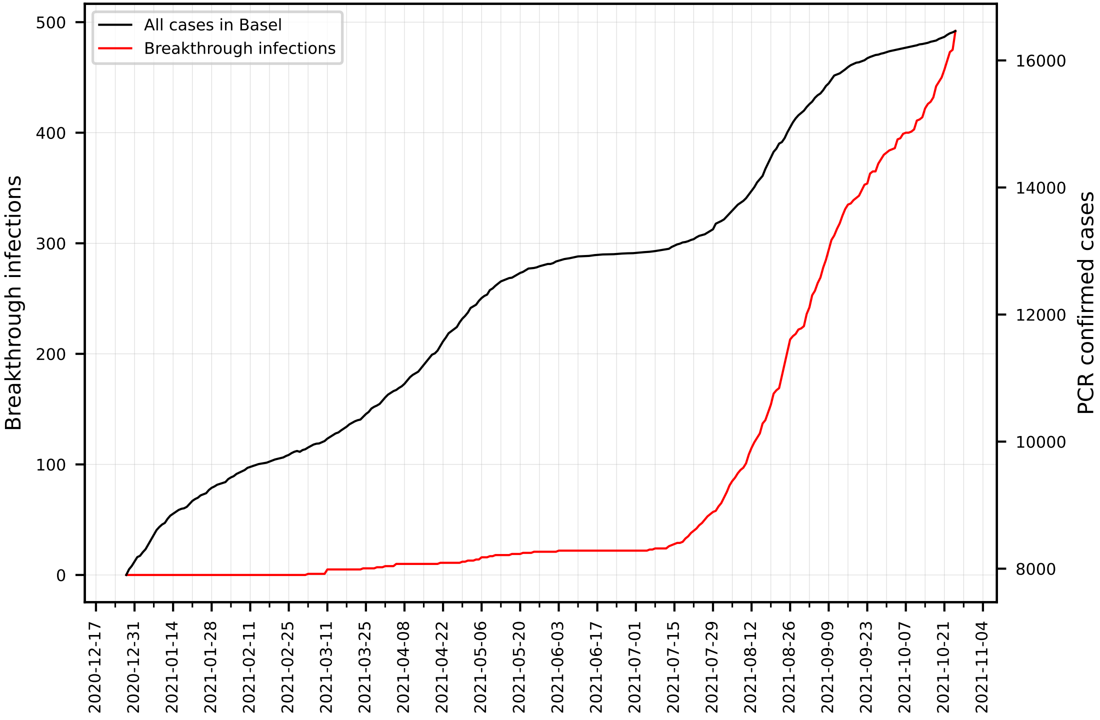

### Supplementary Figure 2A

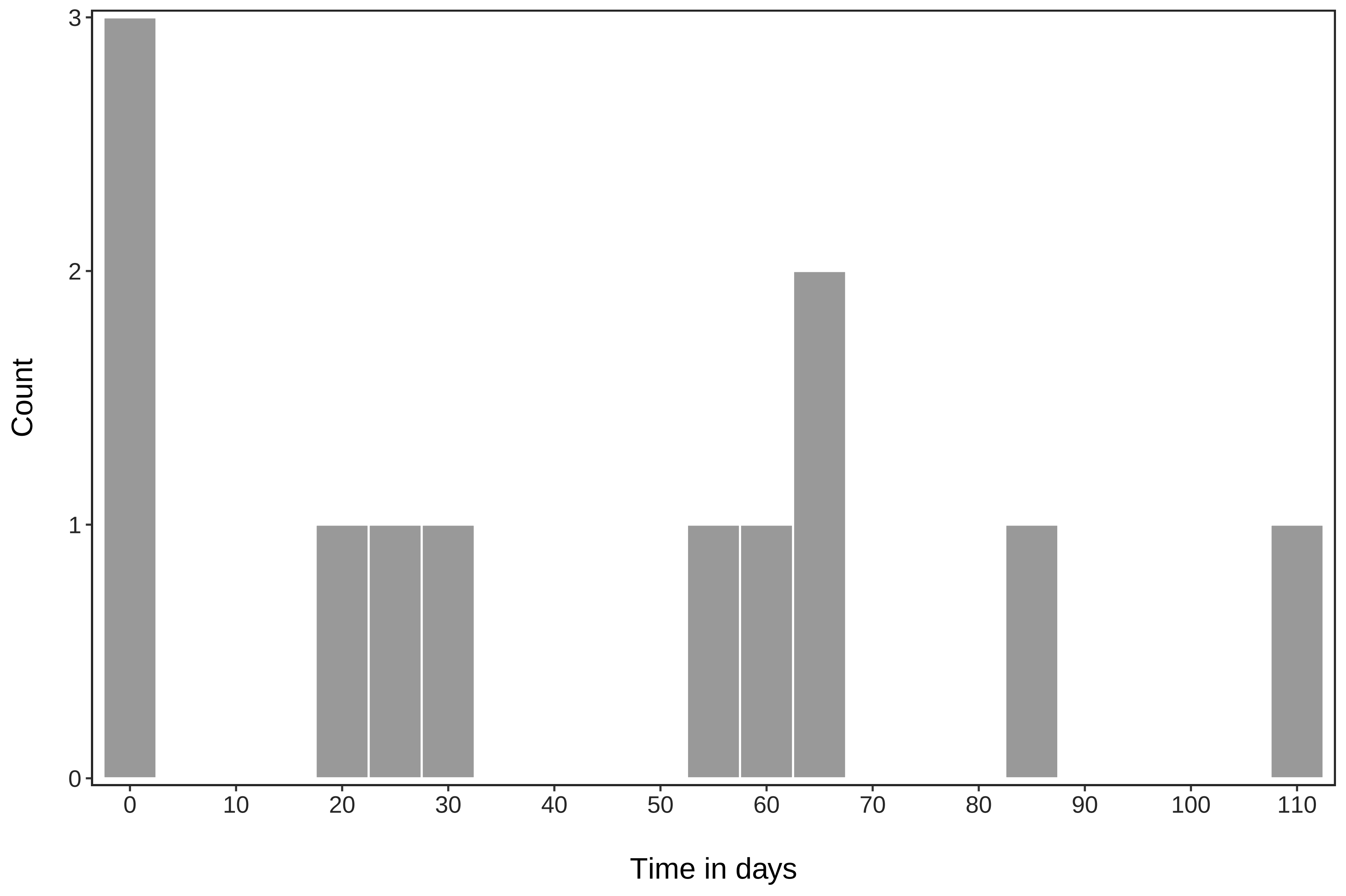

### Supplementary Figure 2B

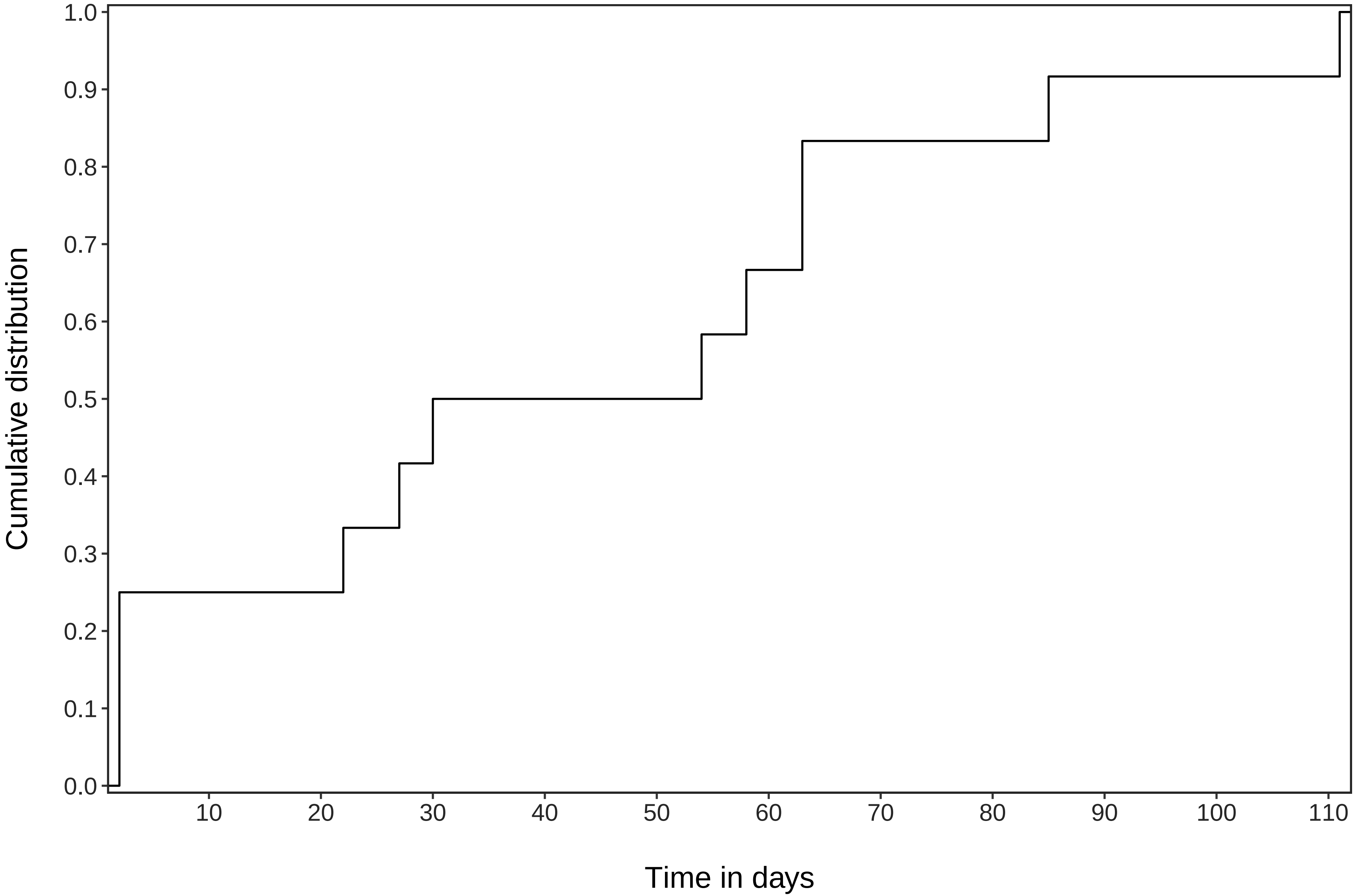

### Supplementary Figure 3A

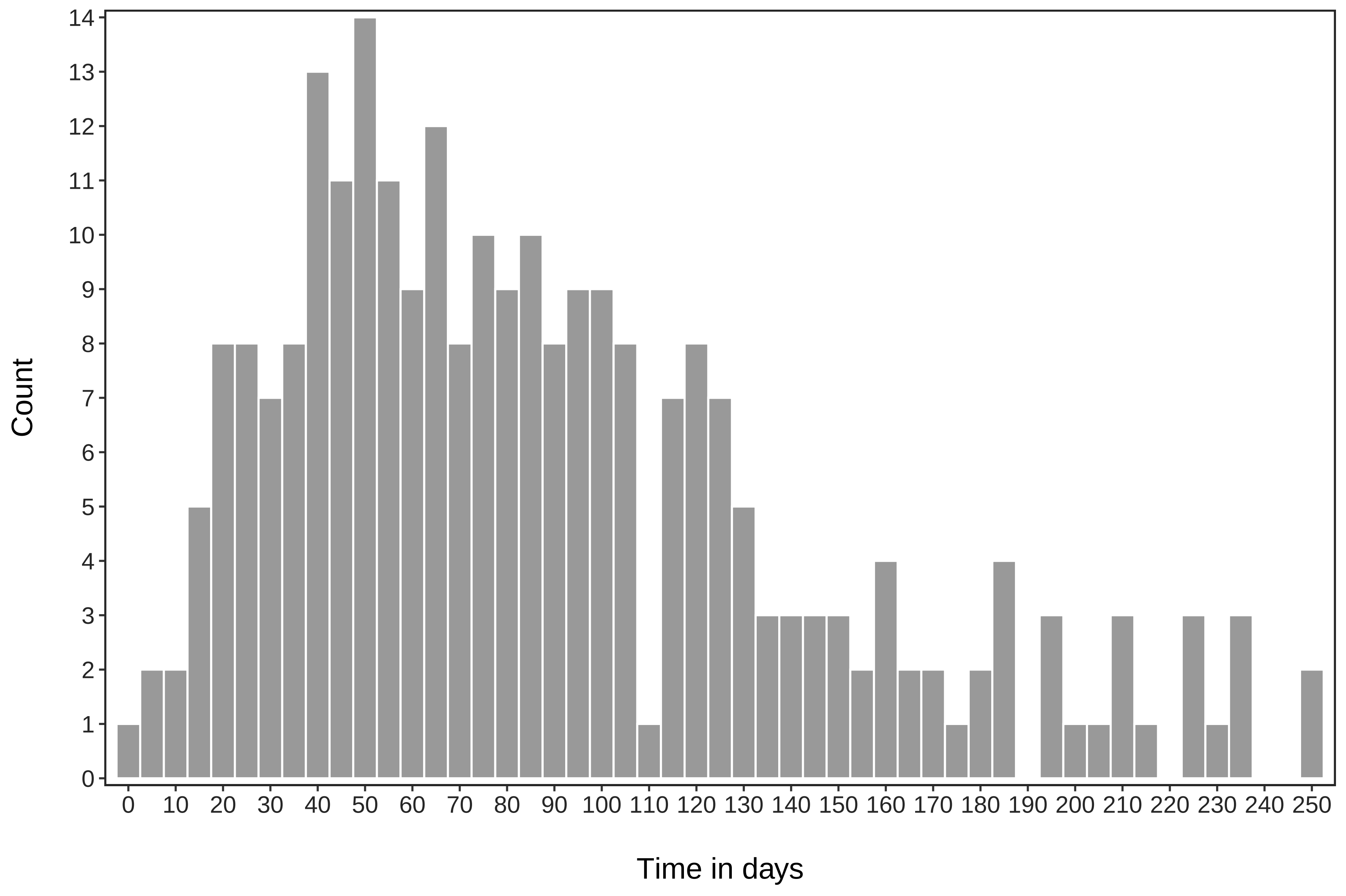

### Supplementary Figure 3B

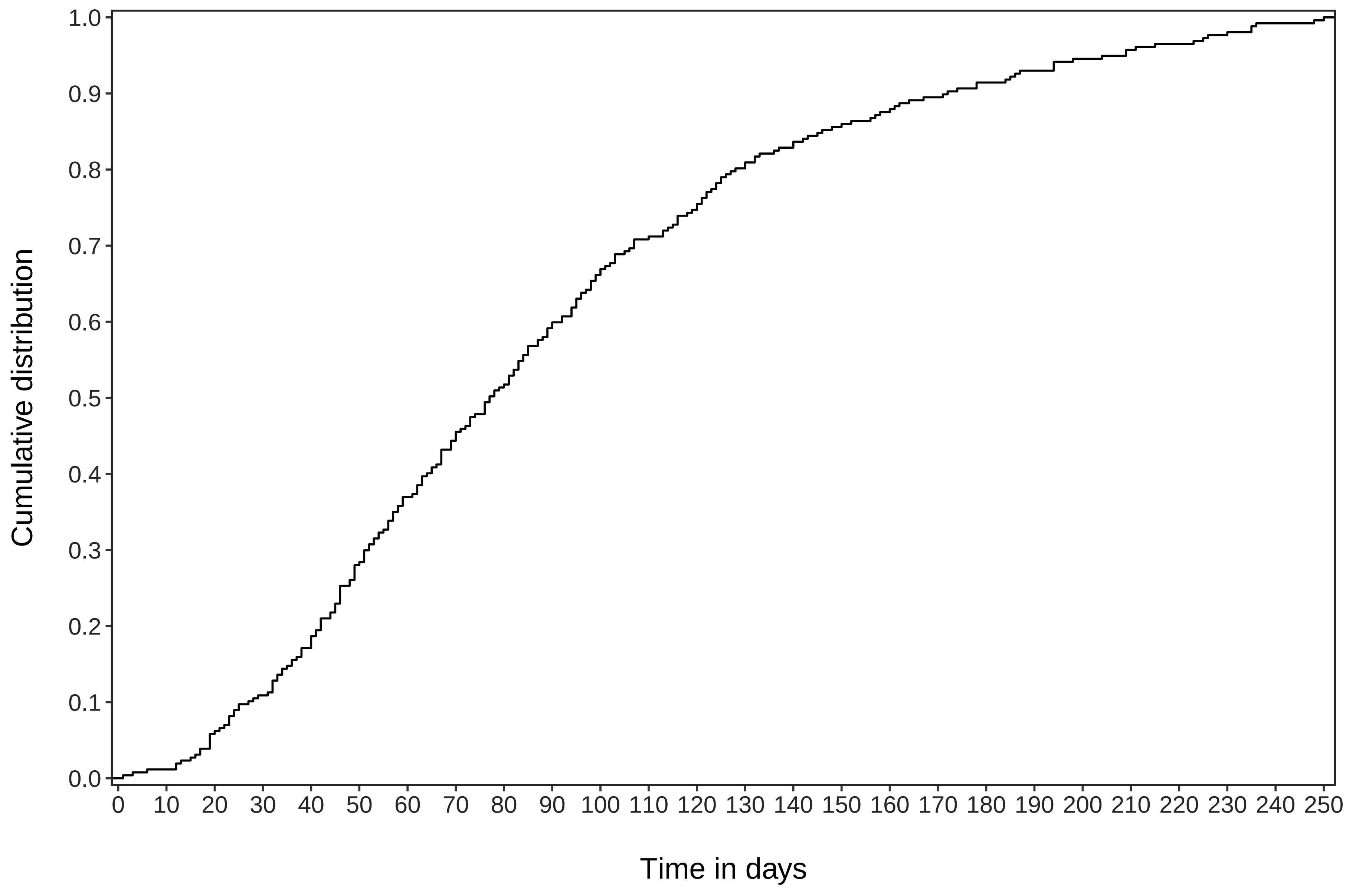
